# Characterizing US contact patterns relevant to respiratory transmission from a pandemic to baseline: Analysis of a large cross-sectional survey

**DOI:** 10.1101/2024.04.26.24306450

**Authors:** Juliana C. Taube, Zachary Susswein, Vittoria Colizza, Shweta Bansal

## Abstract

**Background:** Contact plays a critical role in infectious disease transmission. Characterizing heterogeneity in contact patterns across individuals, time, and space is necessary to inform accurate estimates of transmission risk, particularly to explain superspreading, predict age differences in vulnerability, and inform social distancing policies. Current respiratory disease models often rely on data from the 2008 POLYMOD study conducted in Europe, which is now outdated and potentially unrepresentative of behavior in the US. We seek to understand the variation in contact patterns across time, spatial scales, and demographic and social classifications, and what social behavior looks like at baseline in the absence of an ongoing pandemic.

**Methods:** We analyze spatiotemporal non-household contact patterns across 10.7 million survey responses from June 2020 - April 2021 post-stratified on age and gender to correct for sample representation. To characterize spatiotemporal heterogeneity in respiratory contact patterns at the county-week scale, we use generalized additive models. In the absence of non-pandemic US contact data, we employ a regression approach to estimate baseline contact and address this gap.

**Findings:** Although contact patterns varied over time during the pandemic, contact is relatively stable after controlling for disease. We find that the mean number of non-household contacts is spatially heterogeneous regardless of disease. There is additional heterogeneity across age, gender, race/ethnicity, and contact setting, with mean contact decreasing with age and lower in women. The contacts of White individuals and contacts at work or social events change the most under increased national incidence.

**Interpretation:** We develop the first county-level estimates of non-pandemic contact rates for the US that can fill critical gaps in parameterizing future disease models. Our results identify that spatiotemporal, demographic, and social heterogeneity in contact patterns is highly structured, informing the risk landscape of respiratory infectious disease transmission in the US.

**Funding:** Research reported in this publication was supported by the National Institutes of Health under award number R01GM123007 and R35GM153478 (SB).

**Research in Context:** *Evidence before this study:* We searched Google Scholar for contact data in the US both during and prior to the pandemic published by February 1, 2024, with the search terms “contact patterns”, “social contact data”, “disease-relevant contacts”, “change in contacts pandemic”, “urban rural social contacts,” and “seasonality in contact patterns”. We reviewed the bibliographies of these articles and included known literature not found via these search criteria. We excluded studies using mobility data, focusing on children, or excluding the United States. Previous work has been limited to the state scale or subsets of counties (e.g., focused on a few cities, a single state, or a few counties within a state) rather than all counties in the US.

*Added value of this study:* We contribute the first high-resolution pandemic contact estimates for the US and infer non-pandemic contact patterns at fine spatial and temporal scales. Our results indicate that the number of contacts is fairly stable over time in the absence of major disease, suggesting that the number of contacts is not a primary driver of respiratory infectious disease seasonality in the US. We also identify groups at greatest disease risk due to higher contacts, including younger adults, men, and Hispanic and Black individuals.

*Implications of all the available evidence:* This study demonstrates the importance of incorporating age-specific and spatial heterogeneity of contact patterns into future disease models to build accurate estimates of transmission risk. We demonstrate that temporal variability in contact patterns is insufficient to drive respiratory infectious disease seasonality, that adaptive behaviors in response to disease shift risk along an urban-rural gradient, and that some vulnerable groups are at increased risk of exposure due to contact. We advocate that geographic and social heterogeneity in exposure to disease due to contact patterns be captured more comprehensively for accurate infectious disease predictions and effective and equitable disease mitigation.

## Introduction

Respiratory infectious disease transmission via direct or droplet routes requires close contact. Research over the last two decades has demonstrated that human contact patterns are highly variable between individuals and across geography [1, 2] and highlighted the consequences of this variability for epidemic outcomes and dynamics [3]. Yet, to date, detailed empirical data on contact patterns across the United States have not been available. This lack of data leaves several important gaps in our understanding of the drivers of disease transmission. For example, we’d like to know what factors (e.g., age, season, or location) influence contact patterns across the US and how these factors contribute to variability in infection risk. This information is essential to design targeted interventions and generate accurate estimates of transmission risk across individuals, space, and time. With a new dataset composed of over 10 million survey responses from June 2020 through April 2021, we developed the first detailed characterization of heterogeneities in human contact patterns across the US.

Most infectious disease models assume homogeneous mixing amongst individuals. That is, all individuals have the same contact rate and ability to transmit disease. Homogeneous mixing models have transformed the prediction and control of disease outbreaks (e.g., through *R*_*t*_ estimation [4]) but produce different epidemic dynamics and outcomes than models that incorporate heterogeneities in contact patterns [3, 5]. The POLYMOD study published in 2008 was the first extensive survey to characterize heterogeneities in routine contact patterns relevant to respiratory infectious disease transmission. The data, from eight European countries, suggested that individual contact rates are not homogeneous but rather heavy-tailed and highly assortative by age [1]. These estimates have been used to understand epidemic dynamics, design vaccine strategies, and predict intervention outcomes (e.g., [6–10]). However, the POLYMOD estimates did not capture dynamic behavior in the presence of disease. To fill this gap, researchers conducted the CoMix study during the COVID-19 pandemic to collect contact data contemporaneous with disease across Europe [11, 12]; estimates from the survey were incorporated into forecasting efforts in the UK with mixed results [13]. Notably, both surveys failed to capture fine-scale temporal variability or social heterogeneity in contact and may not be representative of behavior in the United States.

Beyond individual heterogeneity, there are several meaningful dimensions across which contact patterns may vary. Spatial heterogeneity in human behavior plays a vital role in disease dynamics (e.g., [14–17]); if contact patterns exhibit spatial heterogeneity, that might explain observed hotspots of disease burden and/or dynamics of disease spread. Likewise, changes in contact patterns over time may contribute to respiratory infectious disease seasonality (e.g., [18, 19]). Contact may also vary based on surrounding disease transmission as individuals shift their behavior to mitigate risk. These potential heterogeneities in contact patterns profoundly affect our understanding and prediction of epidemic dynamics and our ability to target behavioral interventions. Yet little empirical data exists on them. To identify the dimensions across which contact meaningfully varies, we need high-resolution contact data across geography, time, demography, social classifications, and in the context of disease transmission.

Recent attention has been given to contact patterns in the US, resulting in several studies published over the past few years (in addition to small/indirect earlier studies [20, 21]), which provide insights into variations in contact patterns across multiple dimensions. Breen et al. [22] demonstrate that contact varies between states but are unable to characterize spatial heterogeneity at finer scales in the US, even though other public health-related behaviors have been demonstrated to vary at the county-level [23, 24]. Dorelien et al. [25] use time-use surveys to demonstrate that contact between urban and rural areas may not differ despite the perception that urban inhabitants have more contacts. On temporal variation in contact, one pre-pandemic study found no variation in adult contacts over time [25], while another observed changes from September to May [20]. Many studies find that contact is higher in younger adults, men, and non-White populations, although which race/ethnicity group has the highest contact depends on the study [20,26–28]. Because these past studies are limited in sample size or resolution, they are constrained in their ability to comprehensively characterize heterogeneities in contact patterns across space, time, and social groups.

Now that the US COVID-19 public health emergency has ended, it is vital to characterize contact patterns during the pandemic but also under non-pandemic baseline conditions. In general, pandemic social distancing reduced overall contact (e.g., [29]). However, past work has demonstrated that adherence to pandemic social distancing was heterogeneous across populations, driven in part by health disparities and social inequities that affect the ability of individuals to engage in behavioral interventions [30] and by a patchwork of non-pharmaceutical interventions (e.g., school closures, work from home, gathering bans, mask requirements) implemented at a variety of spatial scales. For example, urban areas exhibited more significant reductions in mobility and, therefore, likely contact [31]. Individuals of higher socioeconomic status are known to have had greater flexibility in their mitigation behavior and could further reduce contacts. Understanding which groups and locations are at the highest infection risk during pandemics and seasonal epidemics is critical for targeted public health surveillance and resource allocation but requires detailed contact data disaggregated by location, age, gender, and race/ethnicity.

Here, we address these pressing gaps by developing fine-scale spatiotemporal estimates of mean non-household contacts in adults. We use an extensive national survey with over 10 million responses collected at the county level and correct for issues of representativeness and small sample size via post-stratification and generalized additive models. We characterize several heterogeneities in contact patterns throughout the pandemic and infer non-pandemic contact patterns by controlling for the effect of disease. We focus on four central questions: (1) How does mean contact vary over time? (2) How does mean contact vary across geography in urban versus rural settings? (3) How do contact patterns vary across age, gender, and race/ethnicity classifications? (4) What are contact patterns like at baseline? Our results are the most comprehensive high-resolution estimates of US contact patterns to date and can inform future disease models in the US, provide insight into local and temporal variation in behavior in response to public health messaging, and contribute to our understanding of drivers of respiratory infectious disease seasonality.

## Methods

In this study, we seek to characterize heterogeneities in non-household contact patterns in the United States using survey data from June 2020 to April 2021. We use survey raking and reweighting of responses to correct for unrepresentative survey sampling. Due to the low sample size in some counties, we use generalized additive models (GAMs) to develop smoothed county-week estimates of the mean number of non-household contacts, disaggregated by geographic, demographic, and social categories. To interrogate whether there is temporal variability in US contact patterns in the absence of a pandemic, we account for the effect of disease incidence on contact with a simple linear regression model and examine the remaining unexplained variability in contact.

### Survey data

We analyzed survey responses of the number of non-household contacts for all 50 US states and the District of Columbia using data from the US COVID-19 Trends and Impact Survey (CTIS) [32]. The CTIS was created by the Delphi Research Group at Carnegie Mellon University and distributed through a partnership with Facebook. Beginning in April 2020, a random state-stratified sample of active Facebook users were invited daily to take the survey about COVID-19 and report how many people they had direct contact with outside their household, where contact was defined as “a conversation lasting more than 5 minutes with a person who is closer than 6 feet away from you, or physical contact like hand-shaking, hugging, or kissing.” Contacts were disaggregated by settings outside the home in the survey question: work, shopping for groceries or other essentials, social gatherings, or other. We analyze the sum of contacts across all these settings starting in June 2020. We removed responses with more than 72 contacts in the last 24 hours (the 95th percentile) and performed sensitivity analyses of this truncation point (appendix p33 to 36). Age, gender, and race were self-reported in the survey, and the survey data are representative of the US population by age, sex, and race/ethnicity (appendix p45). To adjust for unrepresentative sampling at the county-scale, we generated response weights to match county age and sex distributions and post-stratified the data (details in supplement). In order to perform raking with ACS data which is based on sex, we had to assume that sex and gender were equivalent and binary. Due to limited sample size and inappropriate entries, we could not consider responses with genders outside of this binary construct, which we recognize as a limitation. As race/ethnicity is not available throughout the full study period, we cannot include it in the raking procedure for all estimates. To test the sensitivity of our estimates to including race/ethnicity in the raking weights, we compared mean contact estimates at the state-month level for September 2020 - April 2021 (appendix p29). We additionally account for race/ethnicity in the raking weights for any race/ethnicity-specific analyses.

County urbanicity was determined using the NCHS 2013 urban-rural classifications, where 1 indicates large central metropolitan areas, and 6 represents rural non-core areas [33].

### Spatiotemporal contact estimation

To address weekly noise and low sample sizes in the data, we estimated smoothed county-week mean non-household contacts with hierarchical generalized additive models (GAMs). GAMs, which fit arbitrary smooth curves to data [34], are a popular form of regression, especially for time series modeling (e.g., [35, 36]). The `mgcv` package for fitting GAMs has been designed for flexible and efficient estimation with structured time series data [34, 37, 38], accounting for partial observations and dropout, and there is a robust literature on the `mgcv`’s applications in this area.

We use penalized thin-plate splines weighted by sample size with a weekly smooth for each state and a factor smooth for each county within the state, with shared smoothness parameters across counties. Thus, information is shared within states but not between; estimates are penalized to the state average, which may be biased towards urban counties. The model is structured as:

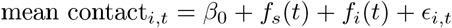

where

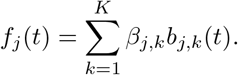

The smooth function *f*_*j*_ is a penalized thin plate regression spline, with *j* as state *s* or county *i* level. [37]. *t* represents each week. The coefficients *β*_*j,k*_ are estimated for each of the *K* basis functions *b*_*j,k*_.

We fit separate GAMs to estimate contact by age, gender, race/ethnicity, and setting at the county or state level for pandemic conditions. Additional details can be found in the supplement.

### Baseline contact estimation

To estimate contact in the absence of a pandemic, we used a linear regression model of weekly contact (as modeled using the GAMs above) predicted by national case incidence [39], state-level policy stringency (measured via the Oxford Stringency Index [40]), county-level policy stringency (calculated as the sum of various ordinal county policies [41–47]), and percentage of the county vaccinated for COVID-19 [24] (appendix p26). We propose that behavioral modification due to risk perception is mediated by disease incidence; to test this, we also considered a subjective measure of risk perception (based on a different question from the CTIS) and find it to be less predictive than incidence (appendix 46). We also considered the role of incidence information across scales (county, state, and national) and found state and national scales to be highly collinear, and county incidence to be less informative relative to national incidence (appendix p46). The Oxford Stringency Index captures the extent of policy limitations on human behavior, including school, work, and transit closures, stay at home requirements, and travel restrictions [40] at the state level. County-level restaurant and bar closures, mask mandates, gathering bans, and stay at home orders compose the county policy variable. (Additional sensitivity analyses related to the policy variables can be found in the appendix p38, 39). We hypothesized that increases in disease incidence would lead to a linear decrease in contact rates, which is supported by the survey data (appendix p24). The model is as follows:

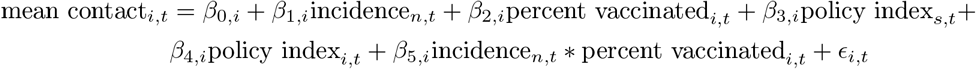

where *i* represents each county, *s* each state, and *n* the nation, *t* represents each week, incidence_*n*_ represents the 4-week rolling average (mean of the previous three weeks and the current week) of national incident cases, policy index_*s*_ represents the Oxford Stringency Index centered at the minimum value observed in the regression period for each state, policy index_*i*_ represents the sum of county level policy metrics centered at the minimum value observed in the regression period for each county, and *ϵ* ~ *N* (0, *σ*^2^). Note that counties are not pooled together which allows us to capture differences in county-level responses to disease and policy metrics. Additional model selection details can be found in the supplement (appendix p46).

Contact after controlling for disease was defined as

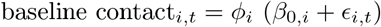

where

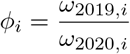

and *!*_*y,i*_ is defined as the mean number of trips into or within a given county *i* for October thru December of year *y*. We added the residual to account for temporal changes in contact not captured by the incidence and policy data. The mobility data is obtained from the SafeGraph Social Distancing dataset [48]. We scale by pre-pandemic mobility data to account for the drastic decrease in contact upon SARS-CoV-2 introduction in the United States. While mobility data do not directly measure contact, they are a reasonable proxy for contact as they are highly correlated (appendix p23).

We fit separate regressions to estimate contact by age, gender, race/ethnicity, and setting at the county or state level for non-pandemic conditions. These models allow us to estimate baseline contact for each demographic/social group by estimating the effect of (overall disease-mediated) risk perception and policy to vary by group.

This analysis was limited to the period from October 2020 to April 2021 to encompass a full “wave” of COVID-19 in the US. Diagnostics for these regression models are provided in the supplement (appendix p29 to 31).

## Results

We characterized heterogeneities in average daily contact patterns at the county-week scale from June 2020 through April 2021 in the United States using 10.7 million valid responses from the CTIS [32] (appendix p19) The survey uses a well-established definition of contact relevant to respiratory disease transmission, and thus offers an advantage over GPS location-based mobility data to characterize contact patterns; past work has also demonstrated that social contact data is more predictive of transmission than mobility data [49–51]. We focus on non-household contacts as they play a crucial role in the dynamics of casual contact infections [52]. To explore variability in contact patterns over time, particularly during the autumn-to-spring period when seasonal changes are expected [53], we developed a statistical model. Rather than capturing all sources of heterogeneity, our model aims to parsimoniously account for pandemic-related effects to estimate non-pandemic contact patterns. We hypothesize that if disease incidence-mediated risk perception and intervention-related behavior change fully explain temporal variability in contact, then baseline behavior remains temporally stable.

### Contact is stable under baseline conditions

Contact varies across the pandemic period, although most counties exhibit similar contact dynamics: increased contact during summer 2020 and spring 2021 and decreased contact during winter 2020-21 (Figure 1A). Some counties, predominantly in Florida, Arizona, Texas, and other parts of the southern US, also reduced their contact in the summer of 2020, coinciding with a COVID-19 surge in the region. This variation appears to be inversely related to SARS-CoV-2 incidence.

**Figure 1.**
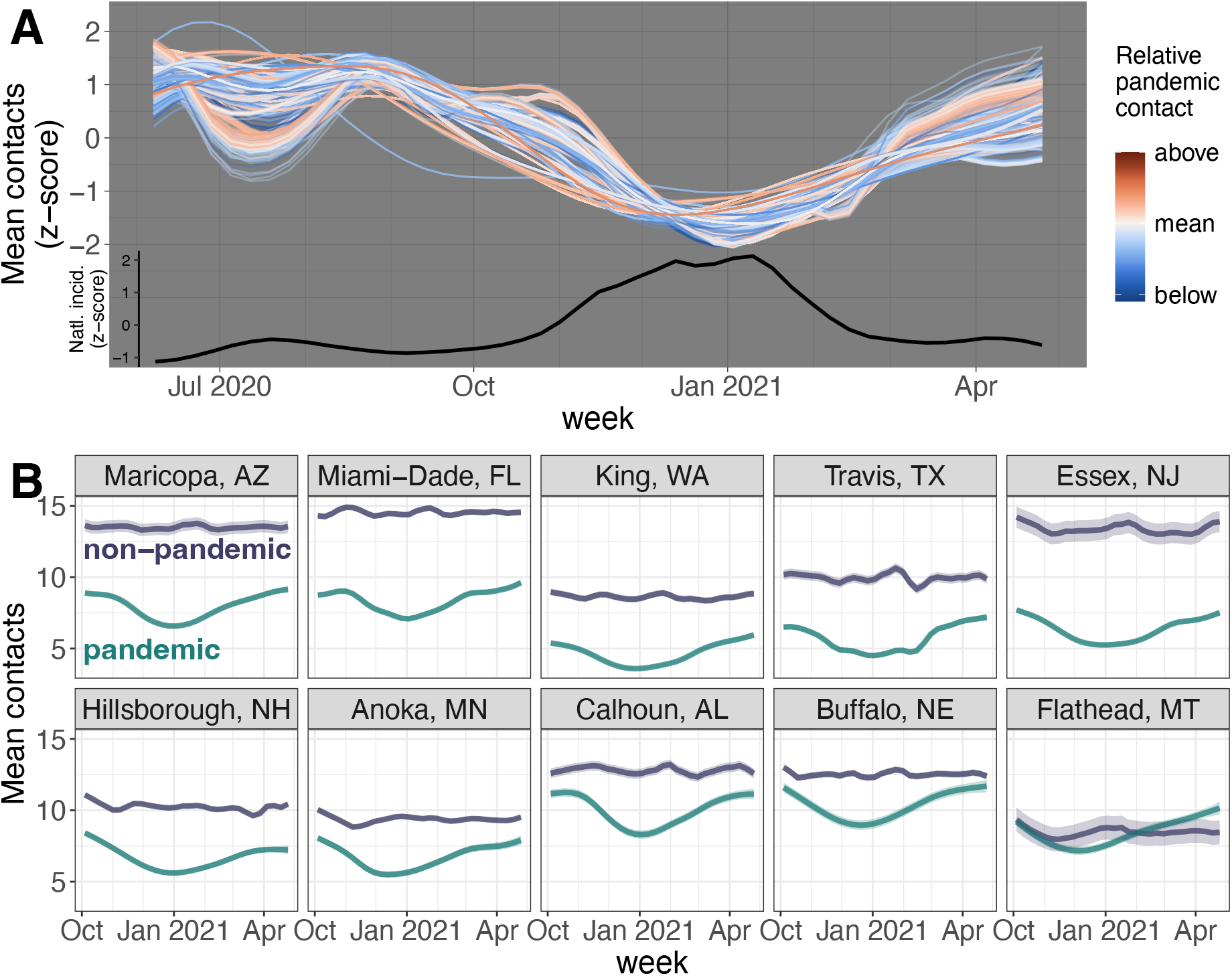
Counties have similar contact dynamics over time and relatively stable contact after controlling for disease. (A) Most counties had higher contact during the summer of 2020, and all had lower contact during the winter of 2020-21. Counties that experienced a dip in contact in the summer of 2020 were typically in states that exhibited higher incidence during that time. Each line represents a county colored by mean contact relative to the national mean (above or below); z-scored contact relative to each county’s mean is shown to allow comparison between time series despite the large range of mean contact values across counties. Black line shows the z-score of centered 3-week rolling average of national case incidence for context. (B) Contact in the absence of disease (slate) is effectively constant over time compared to observed contact during the pandemic (teal) across a diverse set of counties. We controlled for disease using a linear regression predicting contact from national case incidence, state and county policy data, and county vaccination coverage. This analysis is restricted to October 2020 to April 2021 to encompass a full wave of COVID-19. Shaded areas represent one standard error above and below the fitted contact value or estimated non-pandemic value. State abbreviations are as follows: AZ: Arizona, FL: Florida, WA: Washington, TX: Texas, NJ: New Jersey, NH: New Hampshire, MN: Minnesota, AL: Alabama, NE: Nebraska, MT: Montana.

We explore this association using regression models with national case incidence, state and county policies, and county vaccination coverage to predict contact from October 2020 to April 2021. After controlling for the effect of disease on behavior, contact is temporally stable across counties (Figure 1B). Remaining fluctuations are neither substantial nor systematic.

We also validate predicted trends in baseline contact after the Omicron wave by using a different question on the CTIS about contact avoidance (appendix p42, 44). Therefore, our baseline contact estimates can be interpreted as conservative predictions for non-pandemic contact beyond the acute phase of the COVID-19 pandemic.

### Contact is geographically heterogeneous but structured by urbanicity

Mean non-household contact is spatially heterogeneous during the pandemic (Figure 2A) and in the absence of a pandemic (Figure 2B). During the pandemic, the highest degree of contact is observed across the central and southern US while the lowest contact rates are observed along the north Atlantic and western coasts. In contrast, under baseline conditions, we predict a change in the geographic pattern of contact rates.

**Figure 2.**
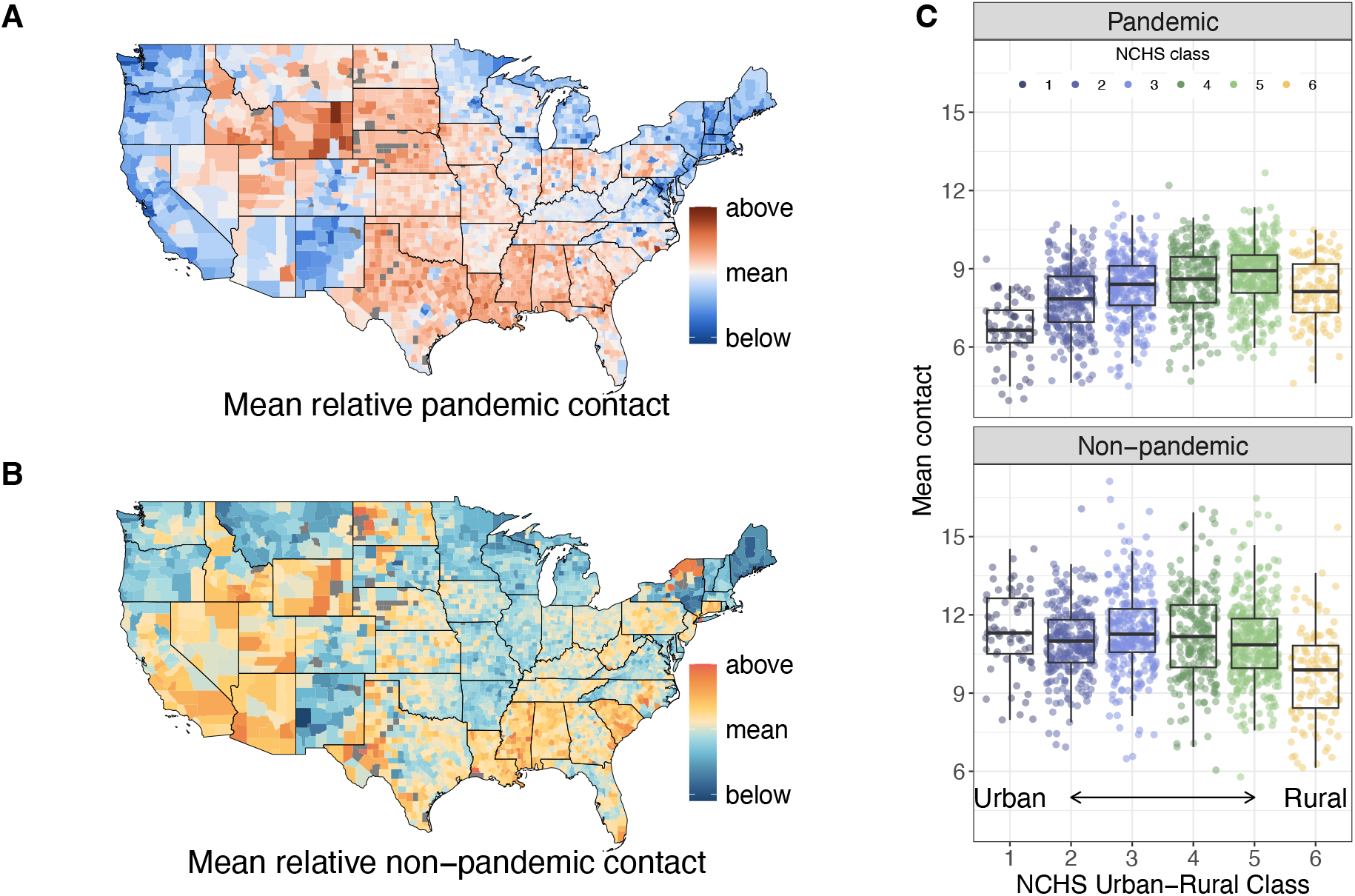
Contact is spatially heterogeneous regardless of disease incidence, but the urban-rural gradient reverses after controlling for disease incidence. (A) Map of mean number of non-household contacts per county relative to the national mean (8.7 contacts) during the pandemic (October 2020 - April 2021). There is high spatial heterogeneity in contact, even within states, which is fairly consistent across time (appendix p28). Gray counties did not have sufficient sample size to estimate contact. (B) Map of inferred mean number of non-household contacts per county relative to the national mean (10.9 contacts) in a non-pandemic scenario. Spatial heterogeneity in contact remains high, though which counties are above and below the national mean has shifted from the pattern observed during the pandemic. (C) Mean number of contacts for each county decreases with increasing urbanicity during the pandemic but increases with urbanicity during inferred non-pandemic times. Only counties with 10 or more responses per week each week (Oct 2020 - Apr 2021) are included. NCHS class describes the urbanicity of the county with 1 representing the most urban large central metro areas, and 6 noncore, rural counties.

We investigate these geographic patterns by considering the association with urbanicity. During the COVID-19 pandemic, respondents in the most urban US counties (NCHS class 1) tended to have fewer contacts compared to more rural counties (Figure 2C). This difference is eliminated by controlling for disease (Figure 2C), suggesting that individuals in urban counties are expected to have slightly more contacts than rural residents under this contact definition in non-pandemic situations.

We demonstrate that these results are robust to truncation in the reported number of contacts (appendix p33 to 36).

### Contact varies demographically and socially

Contact also varies across demographic and social classifications during the pandemic and at baseline. Older respondents tend to have fewer contacts, with individuals between 18 and 54 reporting about the same numbers of contacts on average across the study period (Figure 3A, appendix p40). Men tend to have more contacts than women (Figure 3B), whereas Hispanic respondents have the most contacts compared to Asian respondents who have the fewest (Figure 3C). Most adult contacts occur in work settings, followed by shopping for essentials (Figure 3D). Using separate regression models and county-specific mobility data, we infer baseline contact for each social category. While responsiveness to disease incidence varies within social classifications (appendix p27), the relationship between baseline contact and each social classification remains the same as during the pandemic (Figure 3). Our baseline results are consistent with those from other studies (appendix p3 to 6). During the pandemic, contact estimates are more variable across studies, potentially due to differences in contact definition, survey design, or survey period (appendix p7 to 18).

**Figure 3.**
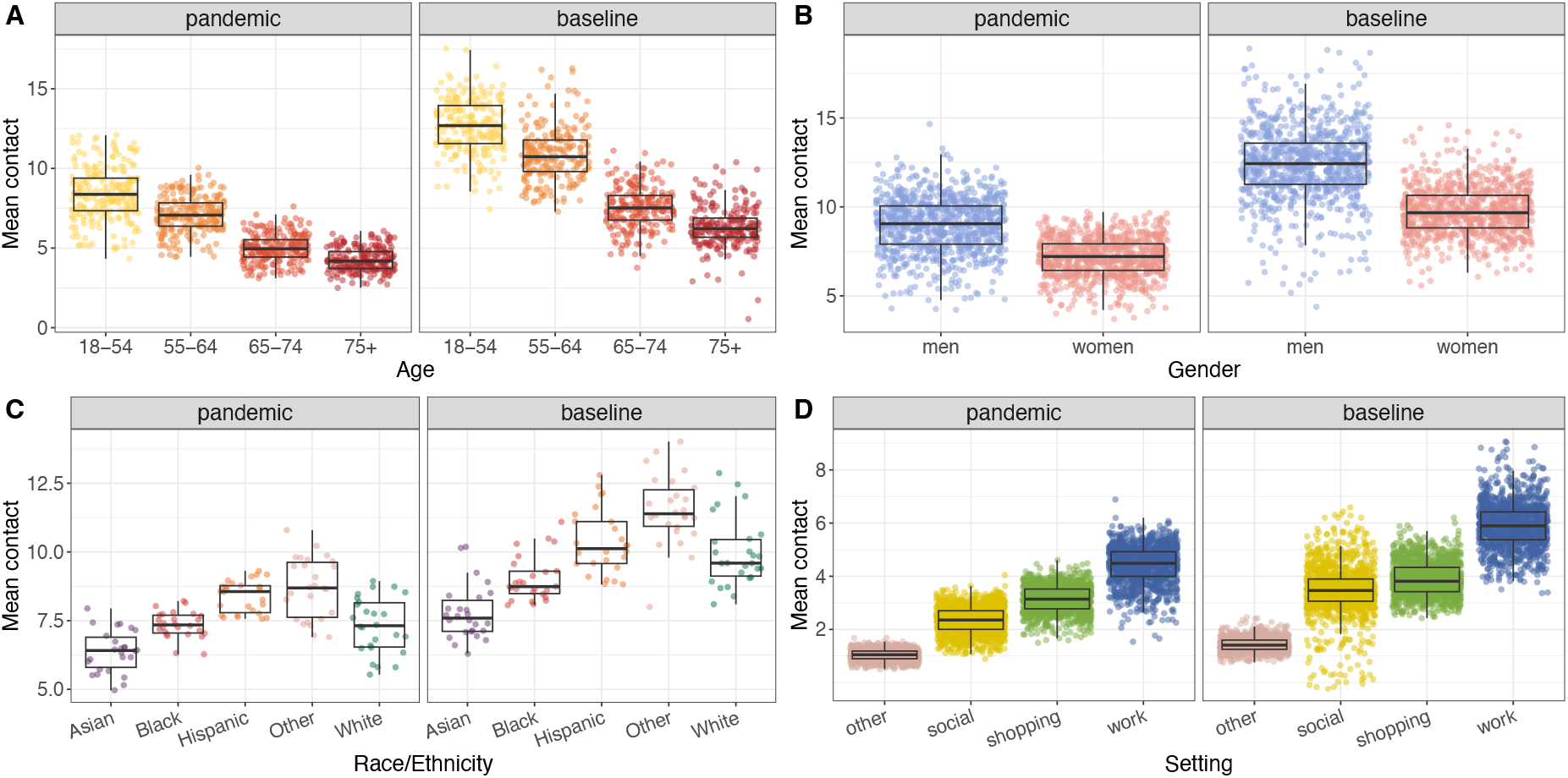
Contact varies across age, gender, race/ethnicity, and setting during the pandemic and at baseline. (A) Mean pandemic and baseline contact by age. Each point represents a county-age category. Analysis is limited to counties with 5+ responses per age category per week. (B) Mean pandemic and baseline contact by gender. Each point represents a county-gender category. Analysis is limited to counties with 5+ responses per gender category per week. (C) Mean pandemic and baseline contact by race/ethnicity. Each point represents a state-race/ethnicity category. Analysis is limited to states with 10+ responses per race/ethnicity category per week. All racial/ethnic categories are non-Hispanic unless labeled otherwise. Other denotes individuals who listed their race as American Indian, Alaska Native, Native Hawaiian, Pacific Islander, or “some other race”. (D) Mean pandemic and baseline contact by setting. Each point represents a county-setting. Analysis is limited to counties with 10+ responses per setting per week.

## Discussion

Interpersonal contact is required for the spread of directly-transmitted pathogens such as SARS-CoV-2. Nevertheless, contact patterns remain poorly understood and difficult to predict. Previous contact studies have focused on European nations and measured contact at coarse spatial and temporal scales (e.g., [1, 22, 28, 54]). These broad scopes leave open questions about how contact patterns vary subnationally, across seasons, and between demographic and social classifications. Here, we estimate non-household contacts at the county-week scale in the US using responses from a large national survey during the COVID-19 pandemic (June 2020 - April 2021). We used post-stratification and generalized additive models weighted by sample size to address sample representation and size issues. We compare our findings to a number of smaller past studies and show consistent patterns. We also use a regression approach to infer non-pandemic contact patterns by controlling for the effect of disease and pandemic interventions. Our findings have several implications for public health researchers and policymakers and can facilitate the much-needed improvement of future disease models and interventions in the US.

We find that most US communities exhibited similar temporal dynamics during the early COVID-19 pandemic which demonstrate an inverse relationship with disease trends. Indeed, after controlling for the effect of incidence-mediated risk perception and disease-related policy, we observe little variability in contact patterns over time suggesting that changes in numbers of contacts cannot explain respiratory infectious disease seasonality. While contact has been demonstrated to differ between summer and winter in other countries [19, 55], our results show that contact does not vary meaningfully from fall to spring, the critical period during which respiratory diseases emerge and fade out in temperate climates. As such, the role of contact in disease seasonality warrants further study; our analysis period was limited to six months which may not capture full annual seasonality, thus longer-term work should investigate whether these trends remain over multiple years and be used to validate our inferences. Importantly, the survey question we use did not differentiate between indoor and outdoor contact or whether individuals were wearing masks; the setting of human contact certainly affects the likelihood of transmission and has been shown to be seasonal [53]. Thus, with the data presented here, we hypothesize that the setting of contact may be a larger driver of seasonality than the number of contacts. Our finding also reinforces other work that fine-grain temporal data may not be necessary for incorporating behavior into infectious disease models [56]. Our hierarchical GAM makes county trajectories within a state more similar by pulling county-level trends toward shared state-level trends, potentially reducing the spatial and temporal variation in our contact estimates.

The high spatial heterogeneity in contact patterns that we observe allows us to identify areas at increased pandemic risk of transmission due to high contact rates, such as the central and southern US. This result also highlights the importance of high-resolution spatial data: there is high variability in contact within states that would be obscured if data are aggregated to the state level. We find that urban counties had fewer contacts on average during the pandemic; this is unsurprising given evidence that urban counties were more responsive to pandemic restrictions [26, 29, 31]. Under baseline conditions, however, we find that urban areas have more contacts on average than suburban or rural areas, which is consistent with behavioral heuristics based on population density. Earlier research has not found a consistent relationship between population density and contact rate, both outside the US [57–59] and in the US [25, 29]. This discrepancy may be explained by differences in contact definition; definitions that are tied to density are likely more representative of aerosol transmission. Given that geographic variation in disease risk creates challenges for disease surveillance, mitigation, and public health communication, further investigation on the role of spatial heterogeneity in behavior is warranted.

Understanding which groups have higher rates of contact is essential for the development of more targeted interventions and to address public health disparities stemming from structural inequities. While the nature of the CTIS precludes any analysis of contact assortativity or clustering, we can identify demographic and social classifications at greater risk due to higher contact rates. Indeed, degree has been shown to be the most important predictor of disease risk compared to other metrics [3]. Like other studies of smaller sample size, both pre-pandemic and pandemic, we find that older adults have fewer contacts than younger adults [1, 27, 28, 60]. However, a limitation of our study (and most other contact studies) is a lack of data on children (individuals aged < 18). We also find that men have more contacts than women during the pandemic, as other national US surveys have shown [27, 28]. Our non-pandemic model shows that this difference persists in baseline conditions, in contrast with [20] (which may be a result of increased contacts by women in the home) and [1] (which finds no meaningful difference in contact between genders). Additionally, we found that Hispanic individuals had the highest contact rates during the pandemic and Asian respondents had the lowest; these results agree with a national survey in 2022 [28] but disagree with pre-pandemic time use data [25]. White respondents showed the most responsiveness to changes in disease incidence, likely reflecting increased ability to work from home. We note, however, that a lack of demographic/social group-specific mobility data may limit our inference of non-pandemic contact patterns across social categories, suggesting potential biases in our baseline contact estimates by social category (appendix p26). Likewise, a lack of incidence data at the county-level disaggregated by demographic group precludes us from analyzing the effect of group-specific incidence on group-specific behavior. Census data show that 93% of US counties only saw a 3% or smaller change in population size from April 2020 to July 2021 [61]. Thus, we expect any changes in the demographic or social distribution within counties due to the pandemic to have negligible effects on the baseline contact estimates. There may be additional biases in the data that we have not addressed, such as social desirability bias, over/under representation by political party, and underrepresentation of rural areas. Overall, our work highlights that social heterogeneities in contact patterns may be responsible for socially structuring transmission risks for respiratory infections and may shape the landscape of response to disease.

In summary, we have developed some of the most detailed pandemic and baseline estimates of contact patterns in the US to date which will be key to informing accurate estimates of transmission risk that account for spatial clustering. Our results can also aid the development of more efficiently targeted interventions. Our work highlights the value of collecting fine-scale behavioral data and the need for long-term longitudinal data collection on contact patterns in the US. We provide some of the first evidence that US adult contact patterns may not vary over time but do vary across counties, with ramifications for understanding respiratory infectious disease seasonality. Improving our understanding of contact patterns, which are such an integral component of disease transmission, should be further prioritized in research efforts going forward.

## Data Availability

Contact estimates, both during the pandemic and at baseline, at the county-week scale and all code to produce and analyze these data are available on GitHub at https://github.com/bansallab/resp_contact. Individual survey responses cannot be shared by the authors, but researchers can refer to https://cmu-delphi.github.io/delphi-epidata/symptom-survey/data-access.html if they would like to enter an agreement for data usage with CMU Delphi.

## Contributors

JCT did the analyses, interpreted the findings, and drafted and edited the manuscript. ZS contributed to the analyses and edited the manuscript. VC interpreted the findings and edited the manuscript. SB conceived and supervised the study, interpreted the findings, and edited the manuscript. JCT, ZS, and SB accessed and verified the data. VC was not part of the data agreement with Carnegie Mellon and so did not have access to the individual survey responses, but did have access to all of the aggregated data. All authors were responsible for the decision to submit the manuscript for publication.

## Data sharing

Contact estimates, both during the pandemic and at baseline, at the county-week scale and all code to produce and analyze these data are available on GitHub at ttps://github.com/bansallab/resp_ contact. Individual survey responses cannot be shared by the authors, but researchers can refer to https://cmu-delphi.github.io/delphi-epidata/symptom-survey/data-access.html if they would like to enter an agreement for data usage with CMU Delphi.

## Acknowledgments

The authors thank the Carnegie Mellon University Delphi team, especially Dr. Alex Reinhart, for sharing the US COVID-19 Trends and Impact Survey results openly and freely and Casey Zipfel for collating pandemic policy data.

## References

1. Mossong J, Hens N, Jit M, Beutels P, Auranen K, Mikolajczyk R, et al. Social Contacts and Mixing Patterns Relevant to the Spread of Infectious Diseases. PLOS Medicine. 2008 Mar;5(3):e74.

2. Hoang T, Coletti P, Melegaro A, Wallinga J, Grijalva CG, Edmunds JW, et al. A Systematic Review of Social Contact Surveys to Inform Transmission Models of Close-contact Infections. Epidemiology. 2019 Sep;30(5):723–736.

3. Bansal S, Grenfell BT, Meyers LA. When Individual Behaviour Matters: Homogeneous and Network Models in Epidemiology. Journal of The Royal Society Interface. 2007;4:879–891.

4. Gostic KM, McGough L, Baskerville EB, Abbott S, Joshi K, Tedijanto C, et al. Practical Considerations for Measuring the Effective Reproductive Number, Rt. PLOS Computational Biology. 2020 Dec;16(12):e1008409.

5. Lloyd-Smith JO, Schreiber SJ, Kopp PE, Getz WM. Superspreading and the Effect of Individual Variation on Disease Emergence. Nature. 2005 Nov;438(7066):355–359.

6. Rohani P, Zhong X, King AA. Contact Network Structure Explains the Changing Epidemiology of Pertussis. Science. 2010 Nov;330(6006):982–985.

7. Mistry D, Litvinova M, Pastore y Piontti A, Chinazzi M, Fumanelli L, Gomes MFC, et al. Inferring High-Resolution Human Mixing Patterns for Disease Modeling. Nature Communications. 2021 Jan;12(1):323.

8. Kretzschmar ME, Rozhnova G, Bootsma MCJ, van Boven M, van de Wijgert JHHM, Bonten MJM. Impact of Delays on Effectiveness of Contact Tracing Strategies for COVID-19: A Modelling Study. The Lancet Public Health. 2020 Aug;5(8):e452–e459.

9. Aleta A, Martín-Corral D, Pastore y Piontti A, Ajelli M, Litvinova M, Chinazzi M, et al. Modelling the Impact of Testing, Contact Tracing and Household Quarantine on Second Waves of COVID-19. Nature Human Behaviour. 2020 Sep;4(9):964–971.

10. Overton CE, Stage HB, Ahmad S, Curran-Sebastian J, Dark P, Das R, et al. Using Statistics and Mathematical Modelling to Understand Infectious Disease Outbreaks: COVID-19 as an Example. Infectious Disease Modelling. 2020 Jan;5:409–441.

11. Gimma A, Munday JD, Wong KLM, Coletti P, van Zandvoort K, Prem K, et al. Changes in Social Contacts in England during the COVID-19 Pandemic between March 2020 and March 2021 as Measured by the CoMix Survey: A Repeated Cross-Sectional Study. PLOS Medicine. 2022 Mar;19(3):e1003907.

12. Coletti P, Wambua J, Gimma A, Willem L, Vercruysse S, Vanhoutte B, et al. CoMix: Comparing Mixing Patterns in the Belgian Population during and after Lockdown. Scientific Reports. 2020 Dec;10(1):21885.

13. Munday JD, Abbott S, Meakin S, Funk S. Evaluating the Use of Social Contact Data to Produce Age-Specific Short-Term Forecasts of SARS-CoV-2 Incidence in England. PLOS Computational Biology. 2023 Sep;19(9):e1011453.

14. Viboud C, Bjørnstad ON, Smith DL, Simonsen L, Miller MA, Grenfell BT. Synchrony, Waves, and Spatial Hierarchies in the Spread of Influenza. Science. 2006 Apr;312(5772):447–451.

15. Favier C, Schmit D, Müller-Graf CDM, Cazelles B, Degallier N, Mondet B, et al. Influence of Spatial Heterogeneity on an Emerging Infectious Disease: The Case of Dengue Epidemics. Proceedings of the Royal Society B: Biological Sciences. 2005 Jun;272(1568):1171–1177.

16. Susswein Z, Valdano E, Brett T, Rohani P, Colizza V, Bansal S. Ignoring Spatial Heterogeneity in Drivers of SARS-CoV-2 Transmission in the US Will Impede Sustained Elimination. medRxiv. 2021 Aug;.

17. Truelove SA, Graham M, Moss WJ, Metcalf CJE, Ferrari MJ, Lessler J. Characterizing the Impact of Spatial Clustering of Susceptibility for Measles Elimination. Vaccine. 2019 Jan;37(5):732–741.

18. Bharti N, Tatem AJ, Ferrari MJ, Grais RF, Djibo A, Grenfell BT. Explaining Seasonal Fluctuations of Measles in Niger Using Nighttime Lights Imagery. Science. 2011 Dec;334(6061):1424–1427.

19. Kummer AG, Zhang J, Jiang C, Litvinova M, Ventura PC, Garcia MA, et al. Evaluating Seasonal Variations in Human Contact Patterns and Their Impact on the Transmission of Respiratory Infectious Diseases. Infectious Diseases (except HIV/AIDS); 2022.

20. DeStefano F, Haber M, Currivan D, Farris T, Burrus B, Stone-Wiggins B, et al. Factors Associated with Social Contacts in Four Communities during the 2007–2008 Influenza Season. Epidemiology & Infection. 2011 Aug;139(8):1181–1190.

21. Zagheni E, Billari FC, Manfredi P, Melegaro A, Mossong J, Edmunds WJ. Using Time-Use Data to Parameterize Models for the Spread of Close-Contact Infectious Diseases. American Journal of Epidemiology. 2008 Nov;168(9):1082–1090.

22. Breen CF, Mahmud AS, Feehan DM. Novel Estimates Reveal Subnational Heterogeneities in Disease-Relevant Contact Patterns in the United States. PLOS Computational Biology. 2022 Dec;18(12):e1010742.

23. Taube JC, Susswein Z, Bansal S. Spatiotemporal Trends in Self-Reported Mask-Wearing Behavior in the United States: Analysis of a Large Cross-sectional Survey. JMIR Public Health and Surveillance. 2023 Mar;9(1):e42128.

24. Tiu A, Susswein Z, Merritt A, Bansal S. Characterizing the Spatiotemporal Heterogeneity of the COVID-19 Vaccination Landscape. American Journal of Epidemiology. 2022 Sep;191(10):1792–1802.

25. Dorélien AM, Ramen A, Swanson I, Hill R. Analyzing the Demographic, Spatial, and Temporal Factors Influencing Social Contact Patterns in U.S. and Implications for Infectious Disease Spread. BMC Infectious Diseases. 2021 Sep;21(1):1009.

26. Dorélien AM, Venkateswaran N, Deng J, Searle K, Enns E, Alarcon Espinoza G, et al. Quantifying Social Contact Patterns in Minnesota during Stay-at-Home Social Distancing Order. BMC Infectious Diseases. 2023 May;23(1):324.

27. Feehan DM, Mahmud AS. Quantifying Population Contact Patterns in the United States during the COVID-19 Pandemic. Nature Communications. 2021 Feb;12(1):893.

28. Nelson KN, Siegler AJ, Sullivan PS, Bradley H, Hall E, Luisi N, et al. Nationally Representative Social Contact Patterns among U.S. Adults, August 2020-April 2021. Epidemics. 2022 Sep;40:100605.

29. Klein B, LaRock T, McCabe S, Torres L, Friedland L, Kos M, et al. Characterizing Collective Physical Distancing in the U.S. during the First Nine Months of the COVID-19 Pandemic. PLOS Digital Health. 2024 Feb;3(2):e0000430.

30. Garnier R, Benetka JR, Kraemer J, Bansal S. Socioeconomic Disparities in Social Distancing During the COVID-19 Pandemic in the United States: Observational Study. Journal of Medical Internet Research. 2021 Jan;23(1):e24591.

31. Zang E, West J, Kim N, Pao C. U.S. Regional Differences in Physical Distancing: Evaluating Racial and Socioeconomic Divides during the COVID-19 Pandemic. PLOS ONE. 2021 Nov;16(11):e0259665.

32. Salomon JA, Reinhart A, Bilinski A, Chua EJ, La Motte-Kerr W, Rönn MM, et al. The US COVID-19 Trends and Impact Survey: Continuous Real-Time Measurement of COVID-19 Symptoms, Risks, Protective Behaviors, Testing, and Vaccination. Proceedings of the National Academy of Sciences. 2021 Dec;118(51):e2111454118.

33. Ingram DD, Franco SJ. 2013 NCHS Urban-Rural Classification Scheme for Counties. Vital and Health Statistics Series 2, Data Evaluation and Methods Research. 2014 Apr;(166):1–73.

34. Wood SN. Generalized Additive Models: An Introduction with R. Second edition ed. Chapman & Hall/CRC Texts in Statistical Science. Boca Raton: CRC Press/Taylor & Francis Group; 2017.

35. Simpson GL. Modelling Palaeoecological Time Series Using Generalised Additive Models. Frontiers in Ecology and Evolution. 2018 Oct;6.

36. Mellor J, Christie R, Overton CE, Paton RS, Leslie R, Tang M, et al. Forecasting Influenza Hospital Admissions within English Sub-Regions Using Hierarchical Generalised Additive Models. Communications Medicine. 2023 Dec;3(1):1–12.

37. Pedersen EJ, Miller DL, Simpson GL, Ross N. Hierarchical Generalized Additive Models in Ecology: An Introduction with Mgcv. PeerJ. 2019 May;7:e6876.

38. Wood SN, L Zheyuan, S Gavin, and Augustin NH. Generalized Additive Models for Gigadata: Modeling the U.K. Black Smoke Network Daily Data. Journal of the American Statistical Association. 2017 Jul;112(519):1199–1210.

39. The New York Times. Coronavirus (Covid-19) Data in the United States; 2021. https://github.com/nytimes/covid-19-data.

40. Hale T, Angrist N, Goldszmidt R, Kira B, Petherick A, Phillips T, et al. A Global Panel Database of Pandemic Policies (Oxford COVID-19 Government Response Tracker). Nature Human Behaviour. 2021 Apr;5(4):529–538.

41. Zipfel C. The Interplay between Human Behavior and Infectious Disease Dynamics. Georgetown University; 2021.

42. Katz R, Toole K, Robertson H, Case A, Kerr J, Robinson-Marshall S, et al. Open Data for COVID-19 Policy Analysis and Mapping. Scientific Data. 2023 Jul;10(1):491.

43. Centers for Disease Control and Prevention. U.S. State and Territorial Gathering Bans: March 11, 2020-August 15, 2021 by County by Day | Data | Centers for Disease Control and Prevention;.

44. Centers for Disease Control and Prevention. U.S. State and Territorial Orders Closing and Reopening Bars Issued from March 11, 2020 through August 15, 2021 by County by Day | Data | Centers for Disease Control and Prevention;.

45. Centers for Disease Control and Prevention. U.S. State and Territorial Orders Closing and Reopening Restaurants Issued from March 11, 2020 through May 31, 2021 by County by Day | Data | Centers for Disease Control and Prevention;.

46. Centers for Disease Control and Prevention. U.S. State and Territorial Public Mask Mandates From April 10, 2020 through August 15, 2021 by County by Day | Data | Centers for Disease Control and Prevention;.

47. Centers for Disease Control and Prevention. U.S. State and Territorial Stay-At-Home Orders: March 15, 2020 – August 15, 2021 by County by Day | Data | Centers for Disease Control and Prevention;.

48. Safegraph. Social Distancing Metrics;. https://docs.safegraph.com/docs/social-distancing-metrics.

49. Crawford FW, Jones SA, Cartter M, Dean SG, Warren JL, Li ZR, et al. Impact of Close Interpersonal Contact on COVID-19 Incidence: Evidence from 1 Year of Mobile Device Data. Science Advances. 2022 Jan;8(1):eabi5499.

50. Tomori DV, Rübsamen N, Berger T, Scholz S, Walde J, Wittenberg I, et al. Individual social contact data and population mobility data as early markers of SARS-CoV-2 transmission dynamics during the first wave in Germanyâan analysis based on the COVIMOD study. BMC medicine. 2021;19:1–13.

51. Delussu F, Tizzoni M, Gauvin L. The Limits of Human Mobility Traces to Predict the Spread of COVID-19: A Transfer Entropy Approach. PNAS Nexus. 2023 Oct;2(10):pgad302.

52. Read JM, Eames KT, Edmunds WJ. Dynamic social networks and the implications for the spread of infectious disease. Journal of The Royal Society Interface. 2008;5(26):1001–1007.

53. Susswein Z, Rest EC, Bansal S. Disentangling the Rhythms of Human Activity in the Built Environment for Airborne Transmission Risk: An Analysis of Large-Scale Mobility Data. eLife. 2023 Apr;12:e80466.

54. CMMID COVID-19 working group, Jarvis CI, Van Zandvoort K, Gimma A, Prem K, Klepac P, et al. Quantifying the Impact of Physical Distance Measures on the Transmission of COVID-19 in the UK. BMC Medicine. 2020 Dec;18(1):124.

55. Nagpal S, Kumar R, Noronha RF, Kumar S, Gupta D, Amarchand R, et al. Seasonal Variations in Social Contact Patterns in a Rural Population in North India: Implications for Pandemic Control. PLOS ONE. 2024 Feb;19(2):e0296483.

56. Pullano G, Alvarez-Zuzek LG, Colizza V, Bansal S. Characterizing US Spatial Connectivity and Implications for Geographical Disease Dynamics and Metapopulation Modeling: Longitudinal Observational Study. JMIR Public Health and Surveillance. 2025 Feb;11(1):e64914.

57. Read JM, Lessler J, Riley S, Wang S, Tan LJ, Kwok KO, et al. Social Mixing Patterns in Rural and Urban Areas of Southern China. Proceedings of the Royal Society B: Biological Sciences. 2014 Jun;281(1785):20140268.

58. Kiti MC, Kinyanjui TM, Koech DC, Munywoki PK, Medley GF, Nokes DJ. Quantifying Age-Related Rates of Social Contact Using Diaries in a Rural Coastal Population of Kenya. PLOS ONE. 2014 Aug;9(8):e104786.

59. Kleynhans J, Tempia S, McMorrow ML, von Gottberg A, Martinson NA, Kahn K, et al. A Cross-Sectional Study Measuring Contact Patterns Using Diaries in an Urban and a Rural Community in South Africa, 2018. BMC Public Health. 2021 Jun;21(1):1055.

60. Mousa A, Winskill P, Watson OJ, Ratmann O, Monod M, Ajelli M, et al. Social Contact Patterns and Implications for Infectious Disease Transmission – a Systematic Review and Meta-Analysis of Contact Surveys. eLife. 2021 Nov;10:e70294.

61. Bureau UC. County Population Totals and Components of Change: 2020-2024; 2025. https://www.census.gov/data/tables/time-series/demo/popest/2020s-counties-total.html.

