## Supplementary materials for "Characterizing US contact patterns relevant to respiratory transmission from a pandemic to baseline: Analysis of a large cross-sectional survey"

### Additional CTIS details

The COVID-19 Trends and Impact Survey was created by researchers in the Delphi Group at Carnegie Mellon University and distributed via Facebook to active users 18 or older starting in April 2020. On a daily basis, a random state-stratified sample of Facebook users are invited to take the survey at the top of their news feed. These users will not be re-invited to take the survey for at least thirty days. The survey asks a broad range of questions related to COVID-19 symptoms and behaviors, with variations across each of the thirteen different waves spanning from April 2020 to June 2022. In this study, we make use of data from Wave 3 (May 21, 2020 - September 7, 2020), Wave 4 (September 8, 2020 - November 23, 2020), Wave 5 (November 24, 2020 - December 18, 2020), Wave 6 (December 19, 2020 - January 11, 2021), Wave 7 (January 12, 2021 - February 7, 2021), Wave 8 (February 8, 2021 - March 1, 2021), and Wave 10 (March 2, 2021 - May 19, 2021; Wave 9 was skipped for numbering purposes).

Weights were provided by Facebook for each response to adjust for non-response and coverage bias at the daily-state level [62]. Briefly, each weight describes the number of people represented by a respondent based on their age, gender, location, and date of response. These weights were calculated via a two-step process using inverse propensity score weighting based on respondents' demographics recorded in their Facebook user profiles to adjust the survey sample to reflect active Facebook users, followed by revisions of these weights using post-stratification so that the survey sample reflects the general population [32]. Because these weights were not at the same scale of our data analysis, we did not use them and instead performed raking to calculate our own weights as described below.

To address the lack of representation in this social media-administered survey, we calculated county-week weights for each observation using the `anesrake` package [64] and the US Census American Community Survey's 2021 data on county age and sex distributions. Age and sex distributions were based on each county's population over 18 to match the survey sample. We use three categories for age (18-24, 25-54, and 55+) and two categories for sex (ACS)/gender (CTIS) (male/man and female/woman). We assume that sex and gender are equivalent because the ACS doesn't collect gender identity information. Subsetting age further results in issues of nonconvergence. Including race or ethnicity was not possible in our raking approach as race/ethnicity data were only available for half the survey period and because convergence was not feasible with it. All analyses explicitly considering race/ethnicity are performed on data raked on race/ethnicity in addition to age and gender, and therefore are at a coarser spatial scale. We estimated mean contact for each county-week using a weighted mean of the responses with the calculated weights. We excluded observations with missing age or gender from the raking process and assigned equal weights to observations from county-weeks that did not converge. Demographic characteristics of respondents before and after weighting and compared to the Census are shown in the appendix, p45.

As part of our data processing, we dropped responses missing fips codes, outside the 50 states and the District of Columbia, and from county-weeks with fewer than three responses.

We focused our analysis on non-household contacts; respondents were not explicitly asked about household contacts, and household contact distributions roughly aligned with those from the Census even without raking on this attribute (appendix p41, 42). We note that non-household and household contacts were not highly correlated at the individual level (Pearson's correlation coefficient = 0.076).

### Spatiotemporal contact estimation details

Generalized additive models were fit with a Gaussian family and identity link using the `bam` function with the `fREML` estimator from the `mgcv` version 1.8-39 R package [34]. To enforce smoother fits for all 50 states, the basis dimensions (k) were set at maximum 30 for the weekly smooth and maximum 15 for the county-level factor smooth with marginal penalty (m) 2 for the weekly smooth and 1 for the county factor smooth, and a gamma value of 2 for each model. For DC, the weekly smooth had a maximum basis dimension of 10 with a penalized cubic regression spline. Diagnostic checks via `gam.check()` and `concurvity()` were completed to ensure that most p-values were not significant, k-indexes were close to 1, k' values were far from each edf, residuals were roughly normally distributed, and there was no heteroscedascity. Model adjusted-$R^2$s varied by state with a mean of 0.44 (appendix p22).

### Baseline contact estimation details & diagnostics

We ran 3,080 linear regression models, one for each county with sufficient data. We show diagnostic plots for a random sample of counties demonstrating that our residuals did not display heteroscedascity or clear trends and are roughly normal (appendix p29 to 31). Across counties, the mean adjusted-$R^2$ was 0.93 and the mean GVIF was 6.00.

### Contact by demographic or social classification model details

#### Age & gender

Using spatiotemporal GAMs, we estimated mean contact in four age groups for each county-week (18-54, 55-64, 65-74, 75+). We had to collapse age groups to have sufficient sample size, but note that contact rates among the 18-54 age group were quite similar prior to aggregation (appendix p40). This analysis was limited to counties with 5 or more responses in each age group for each of the 30 weeks from October 2020 through April 2021 (224 counties). For each age group separately, we estimated pandemic contact using the spatiotemporal GAMs and baseline contact using linear regression as described above. The same steps were followed for the two gender classes in the survey; there were 889 counties with sufficient sample size to estimate contact by gender. Given these sample size constraints, these results may not be representative of differences in contact patterns between age (and possibly gender) groups in the most rural US counties. This setup does allow us to compare age- and gender-specific regression coefficients describing the association between contact and disease incidence for each age/gender class.

#### Race/ethnicity

Using spatiotemporal GAMs, we estimated mean contact in five race/ethnicity categories (Asian, Black, Hispanic, other, and White) for each state-week. This analysis was limited to states with 10 or more responses in each racial/ethnic category for each of the 30 weeks from October 2020 through April 2021 (24 states) and was performed on the data that was reweighted by race/ethnicity, in addition to age and gender. Due to limited sample size, this analysis had to be performed at the state instead of the county level. For each racial/ethnic category separately, we estimated pandemic contact using the spatiotemporal GAMs and baseline contact using linear regression as described above with a couple exceptions. In the original GAMs we allowed for information sharing between counties within the same state; for these race/ethnicity-specific GAMs we instead allow for information sharing between states in the same census region of the US (West, Midwest, Northeast, South). For the linear regression, we predict contact using national incidence, state vaccination coverage, and state Oxford Stringency Index.

#### Setting

Using spatiotemporal GAMs, we estimated mean contact in four settings (work, social gatherings, shopping for essentials, and other) for each county-week. This analysis was limited to counties with 10 or more responses in each setting for each of the 30 weeks from October 2020 through April 2021. For each setting separately, we estimated pandemic contact using the spatiotemporal GAMs and baseline contact using linear regression as described above.

### Comparison to existing contact studies

We validated our findings with other work both before and during the pandemic, in appendix p3 to 18 below.

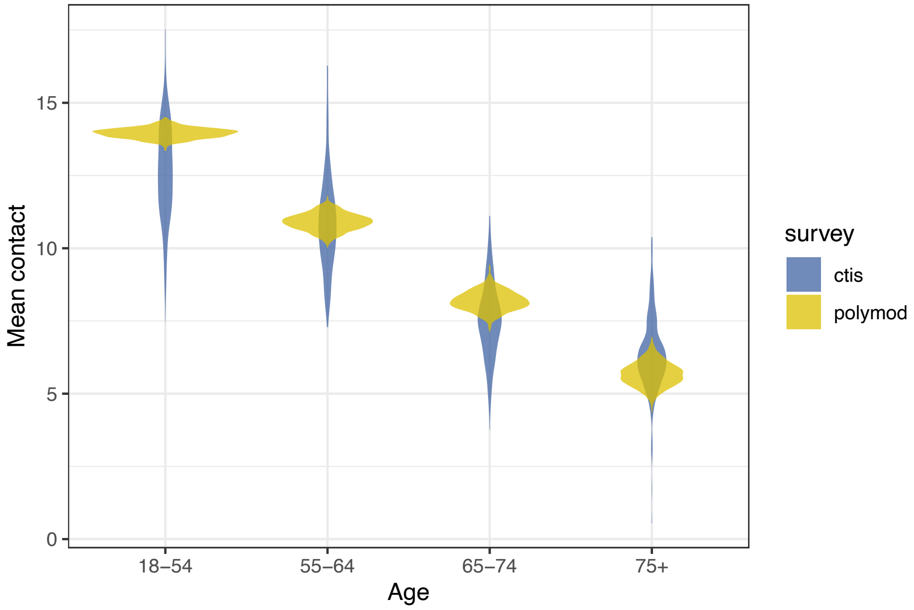


**Figure S1. Comparison between our baseline contact estimates by age with those from the POLYMOD study [1].** Yellow violins are composed of bootstrapped means from 1,000 samples from the POLYMOD study (all responses equally weighted, truncated at 72 contacts, over all countries). Blue violins show the distribution of mean county-age group baseline estimates inferred from the CTIS.

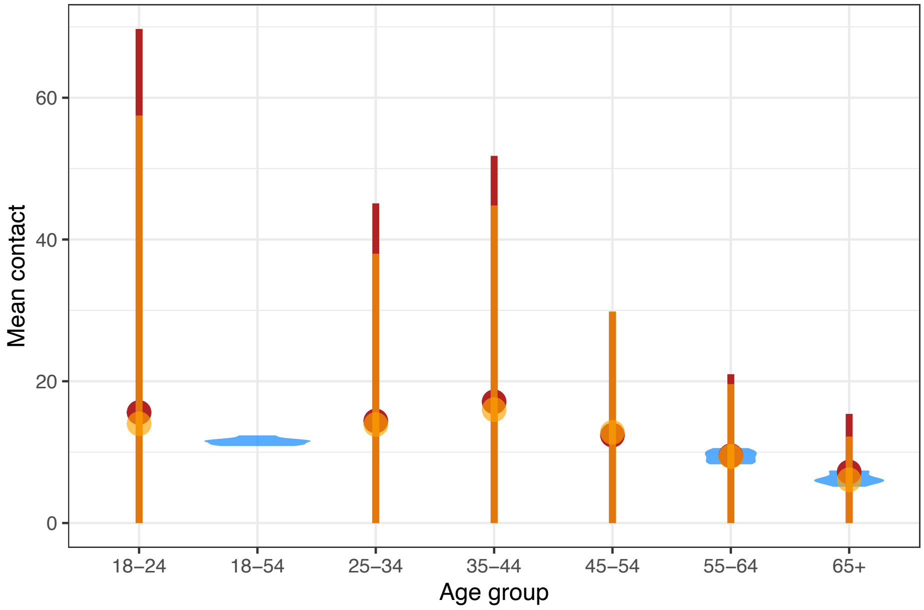


**Figure S2. Comparison between our baseline contact estimates by age with those from [20].** Destefano and colleagues' [20] analyzed contact patterns across four counties in North Carolina, USA during the 2007-08 flu season, with 3,845 survey respondents. They measured both the mean number of speaking interactions (orange), defined as face to face conversations lasting at least one minute, and mean number of close proximity contacts (red), defined as within 6 feet for at least fifteen minutes. Point ranges represent one standard deviation above and below the mean (truncated at 0). Blue violins show the distribution of mean county-age group baseline estimates for seven North Carolina counties (those with sufficient group sample size) inferred from the CTIS.

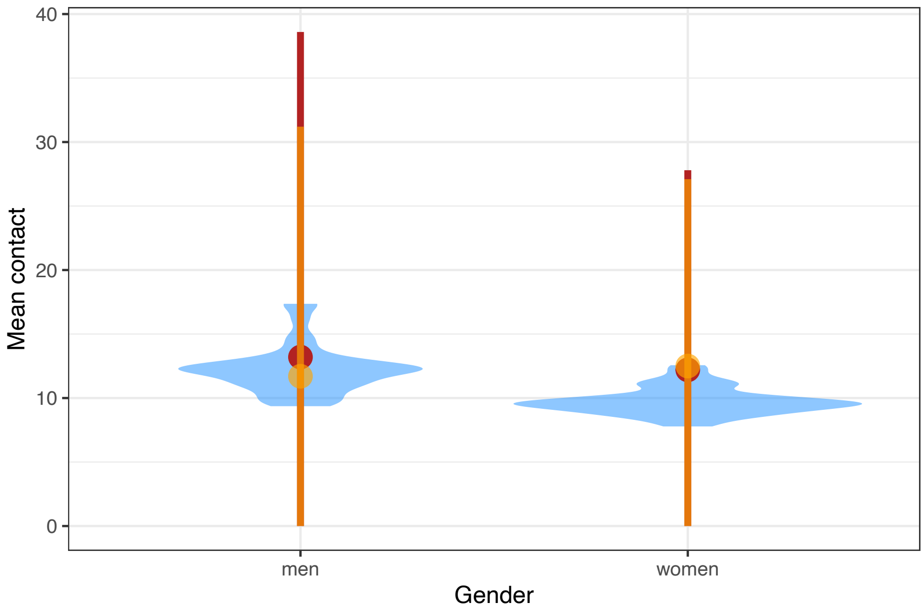


**Figure S3. Comparison between our baseline contact estimates by gender with those from [20].** Destefano and colleagues' [20] analyzed contact patterns across four counties in North Carolina, USA during the 2007-08 flu season, with 3,845 survey respondents. They measured both the mean number of speaking interactions (orange), defined as face to face conversations lasting at least one minute, and mean number of close proximity contacts (red), defined as within 6 feet for at least fifteen minutes. Point ranges represent one standard deviation above and below the mean (truncated at 0). Blue violins show distribution of mean county-gender baseline estimates for 36 North Carolina counties (those with sufficient group sample size) inferred from the CTIS.

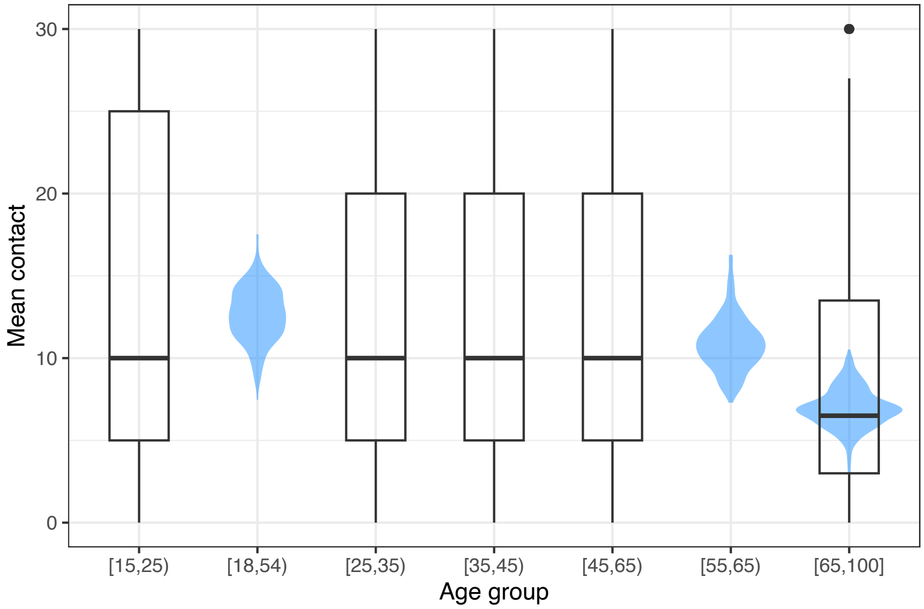


**Figure S4. Comparison between our baseline contact estimates by age with those from [68].** Feehan and Cobb [68] conducted a national study of 3,076 Facebook users 15 years of age and older in the US before the pandemic (2015). They defined a contact as a face to face conversation containing at least three words. Boxplots were constructed using the raw data downloaded from `https://doi.org/10.7910/DVN/M74AJ4` `fb_ego.tab` file, where the center black line represents the median, the box summarizes the middle 50th percentile, the whiskers show the 1.5 times the interquartile range, and dots represent outliers. There are 14 outliers all with the same value of 30 reported contacts so the single dot above the [65,100] box is composed of 14 dots all on top of one another. Blue violins show distribution of mean county-age group baseline estimates inferred from the CTIS.

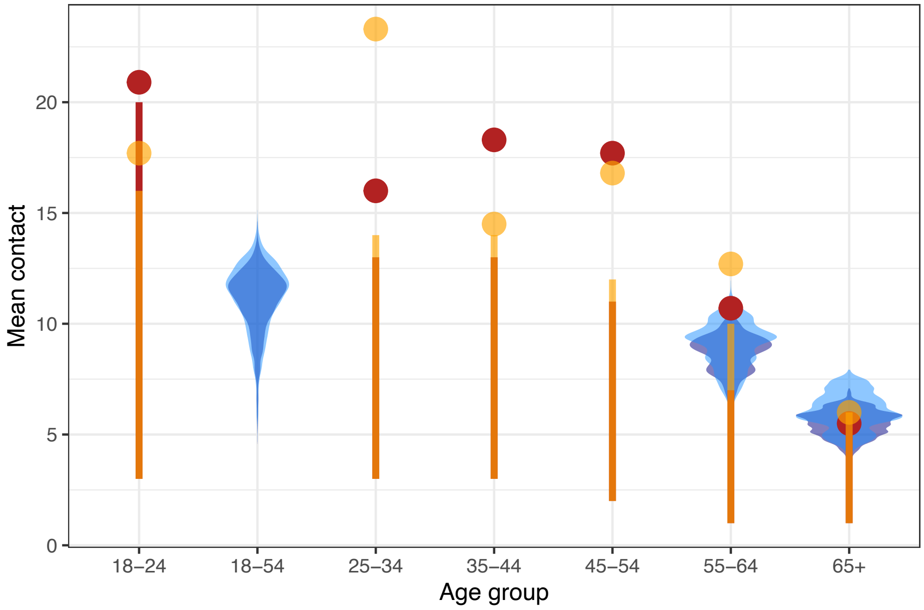


**Figure S5. Comparison between our pandemic contact estimates by age with those from [28].** Nelson and colleagues' [28] conducted a nationally representative study of 3,112 individuals in the US during two periods, August - December 2020 (red) and March - April 2021 (orange). Contacts were defined as interactions within 6 feet involving the exchange of at least 3 words or involving physical touch. Red and orange points show the mean number of contacts for each time period, while ranges stretch from the 25th to 75th percentiles for each time period. Note that [28] truncated contacts at 50 per day and had smaller age group bins. Blue violins show the distribution of mean county-age group estimates from the CTIS: dark blue for August - December 2020 and light blue for March - April 2021 to match the time periods in Nelson et al., i.e. dark red and dark blue plots cover similar time periods and light orange and light blue plots cover similar time periods.

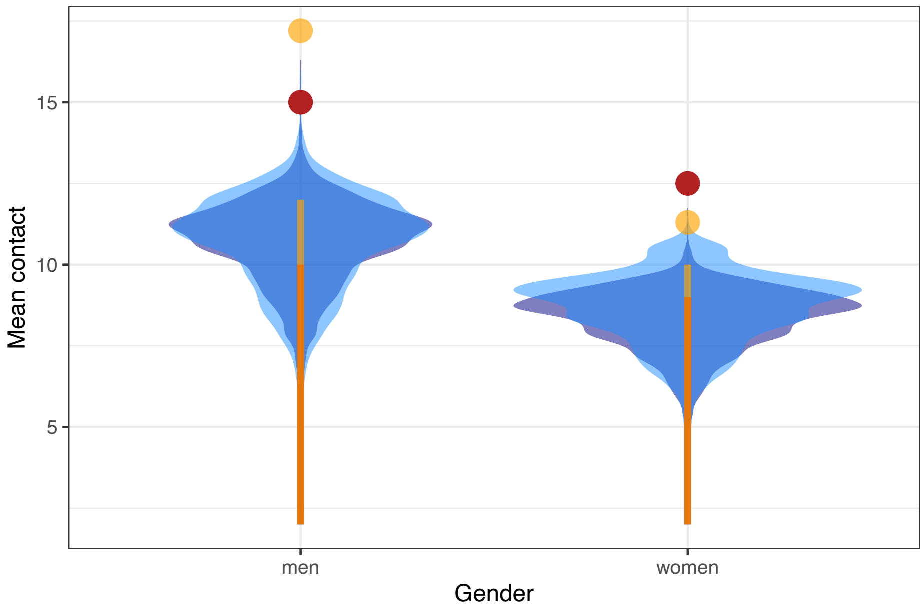


**Figure S6. Comparison between our pandemic contact estimates by gender with those from [28].** Nelson and colleagues' [28] conducted a nationally representative study of 3,112 individuals in the US during two periods, August - December 2020 (red) and March - April 2021 (orange). Contacts were defined as interactions within 6 feet involving the exchange of at least 3 words or involving physical touch. Points show the mean number of contacts while ranges stretch from the 25th to 75th percentiles. Note that [28] truncated contacts at 50 per day. Blue violins show distribution of mean county-gender group estimates from the CTIS: dark blue for August - December 2020 and light blue for March - April 2021 to match the time periods in Nelson et al., i.e. dark red and dark blue plots cover similar time periods and light orange and light blue plots cover similar time periods.

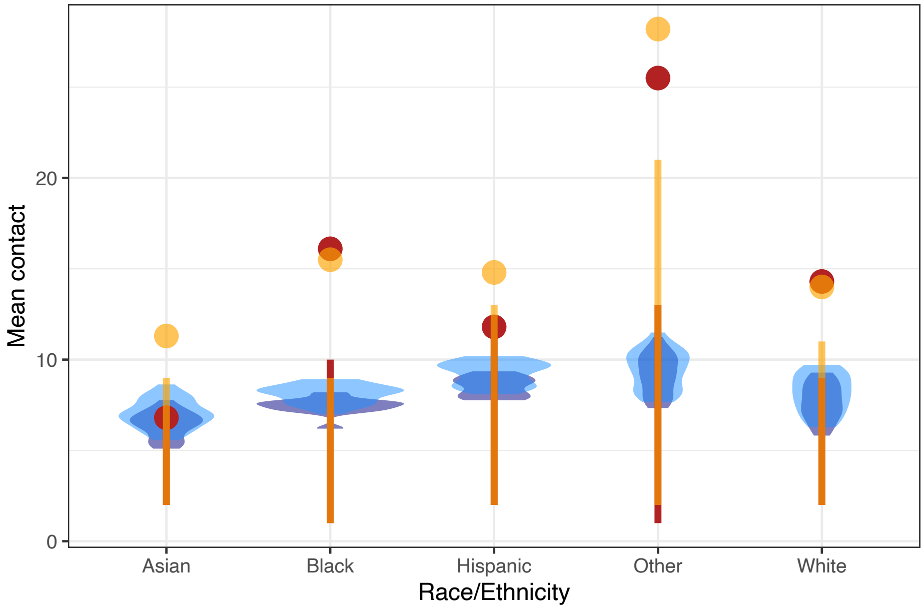


**Figure S7. Comparison between our pandemic contact estimates by race/ethnicity with those from [28].** Nelson and colleagues' [28] conducted a nationally representative study of 3,112 individuals in the US during two periods, August - December 2020 (red) and March - April 2021 (orange). Contacts were defined as interactions within 6 feet involving the exchange of at least 3 words or involving physical touch. Points show the mean number of contacts while ranges stretch from the 25th to 75th percentiles. Note that [28] truncated contacts at 50 per day. Blue violins show distribution of mean state-race/ethnicity group estimates from the CTIS: dark blue for September - December 2020 and light blue for March - April 2021 to match the time periods in Nelson et al., i.e. dark red and dark blue plots cover similar time periods and light orange and light blue plots cover similar time periods.

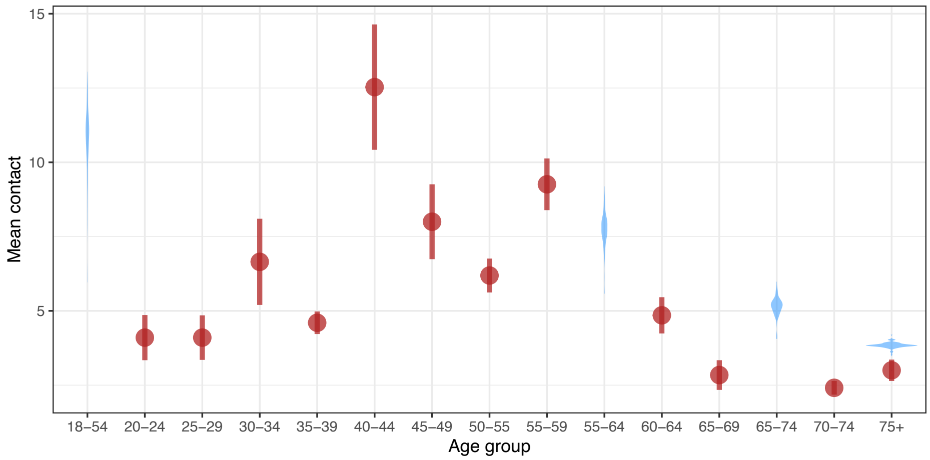


**Figure S8. Comparison between our pandemic contact estimates by age with those from [26].** Dorelien and colleagues' [26] measured contact patterns in 2,083 children and adults in Minnesota, USA in April and May 2020. Contacts were defined as two-way conversations with 3+ words in the physical presence of another person or physical skin-to-skin contact. Red point ranges denote mean contact and one standard deviation above and below the mean from uncensored contact data. Blue violins show distribution of mean county-age group estimates for all 87 Minnesota counties from the CTIS for the first two weeks of May 2020 (these data are available in the CTIS but not as reliable).

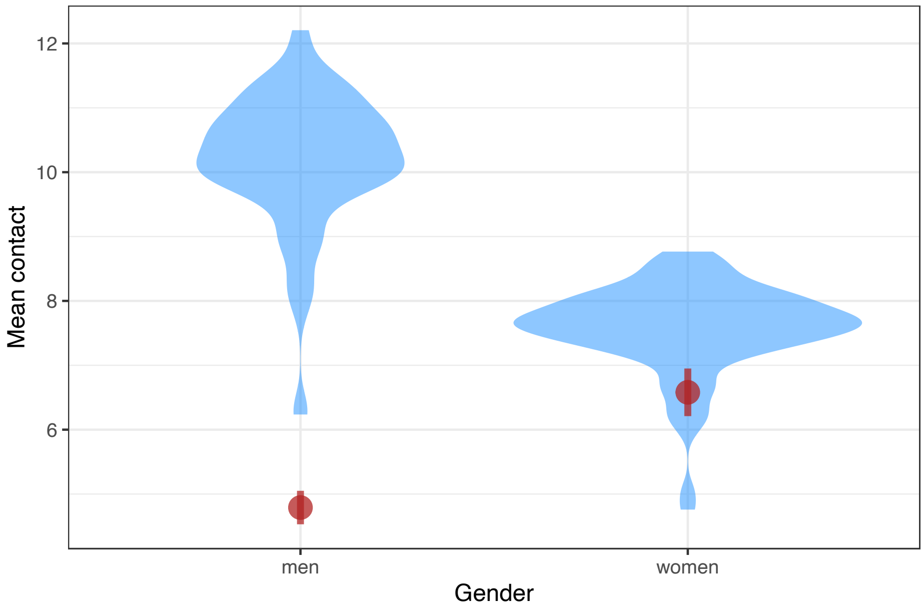


**Figure S9. Comparison between our pandemic contact estimates by gender with those from [26].** Dorelien and colleagues' [26] measured contact patterns in 2,083 children and adults in Minnesota, USA in April and May 2020. Contacts were defined as two-way conversations with 3+ words in the physical presence of another person or physical skin-to-skin contact. Red point ranges denote mean contact and one standard deviation above and below the mean from uncensored contact data. Blue violins show distribution of mean county-gender group estimates for all 87 Minnesota counties from the CTIS for the first two weeks of May 2020 (these data are available in the CTIS but not as reliable).

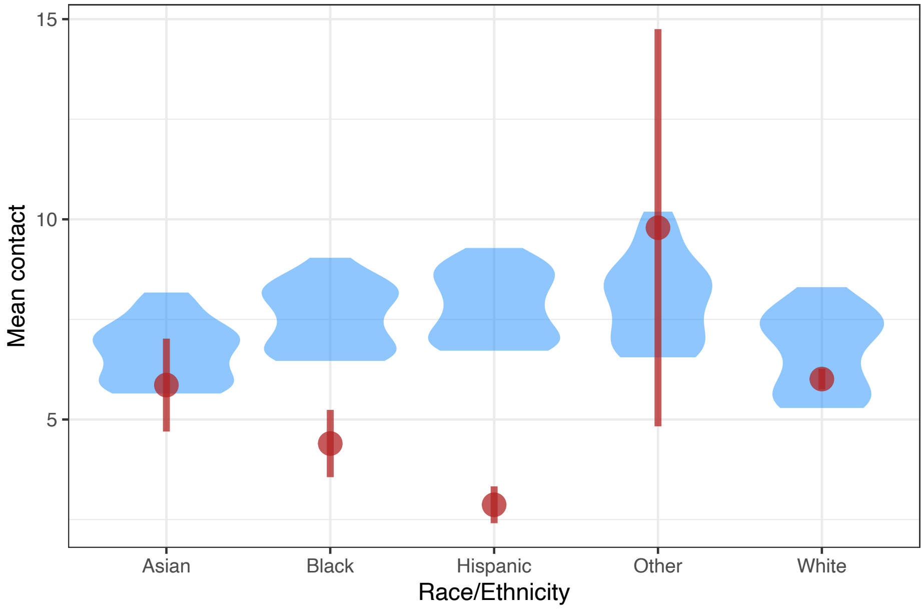


**Figure S10. Comparison between our pandemic contact estimates by race/ethnicity with those from [26].** Dorelien and colleagues' [26] measured contact patterns in 2,083 children and adults in Minnesota, USA in April and May 2020. Contacts were defined as two-way conversations with 3+ words in the physical presence of another person or physical skin-to-skin contact. Red point ranges denote mean contact and one standard deviation above and below the mean from uncensored contact data. Blue violins show distribution of mean Minnesota race/ethnicity estimates from the CTIS for each week from September 2020 to April 2021 (a non-overlapping time period).

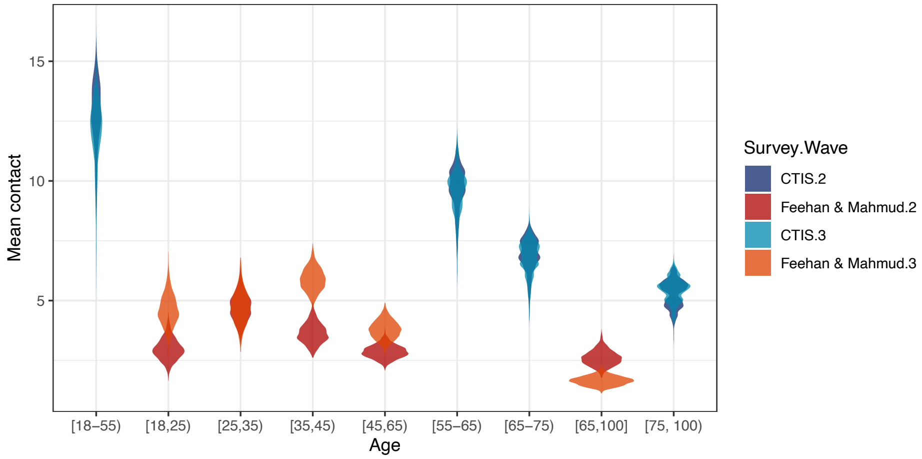


**Figure S11. Comparison between our pandemic contact estimates by age with those from [27].** Feehan and Mahmud [27] conducted a nationally representative survey of contact patterns in the US including one week in June (Wave 2) and one week in September 2020 (Wave 3). Across the two waves, they had 2,641 respondents who reported fewer than 72 contacts. Red and orange violins show the bootstrapped mean contact for June and September 2020, respectively, using provided weights from [27]. (Note that we have imposed a truncation point on their data to match our analysis so the weights may no longer be accurate.) Blue and teal violins show the distribution of mean county-age group estimates from the CTIS for the same weeks in June and September, respectively.

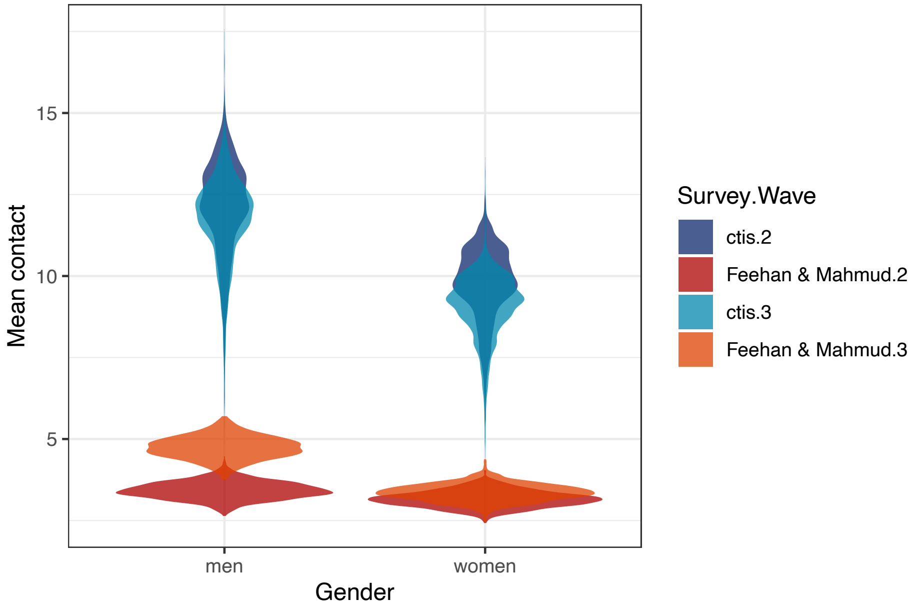


**Figure S12. Comparison between our pandemic contact estimates by gender with those from [27].** Feehan and Mahmud [27] conducted a nationally representative survey of contact patterns in the US including one week in June (Wave 2) and one week in September 2020 (Wave 3). Across the two waves, they had 2,641 respondents who reported fewer than 72 contacts. Red and orange violins show the bootstrapped mean contact for June and September 2020, respectively, using provided weights from [27]. (Note that we have imposed a truncation point on their data to match our analysis so the weights may no longer be accurate.) Blue and teal violins show the distribution of mean county-gender estimates from the CTIS for same weeks in June and September, respectively.

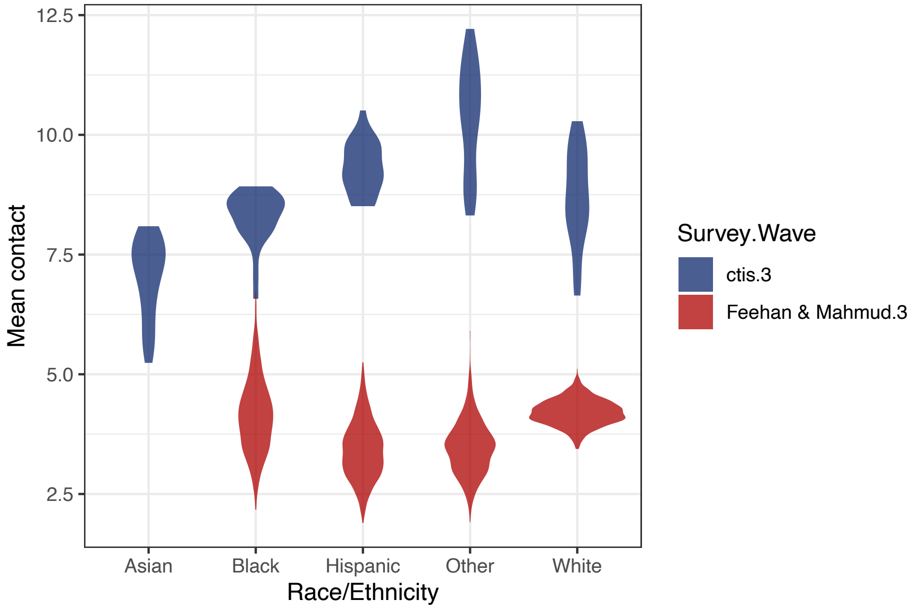


**Figure S13. Comparison between our pandemic contact estimates by race/ethnicity with those from [27].** Feehan and Mahmud [27] conducted a nationally representative survey of contact patterns in the US including one week in September 2020 (Wave 3). In this wave, they had 1,514 respondents who reported fewer than 72 contacts. Red violins show the bootstrapped mean contact for September 2020, using provided weights from [27]. (Note that we have imposed a truncation point on their data to match our analysis so the weights may no longer be accurate.) Blue violins show distribution of mean state-race/ethnicity estimates from the CTIS for the same week in September 2020.

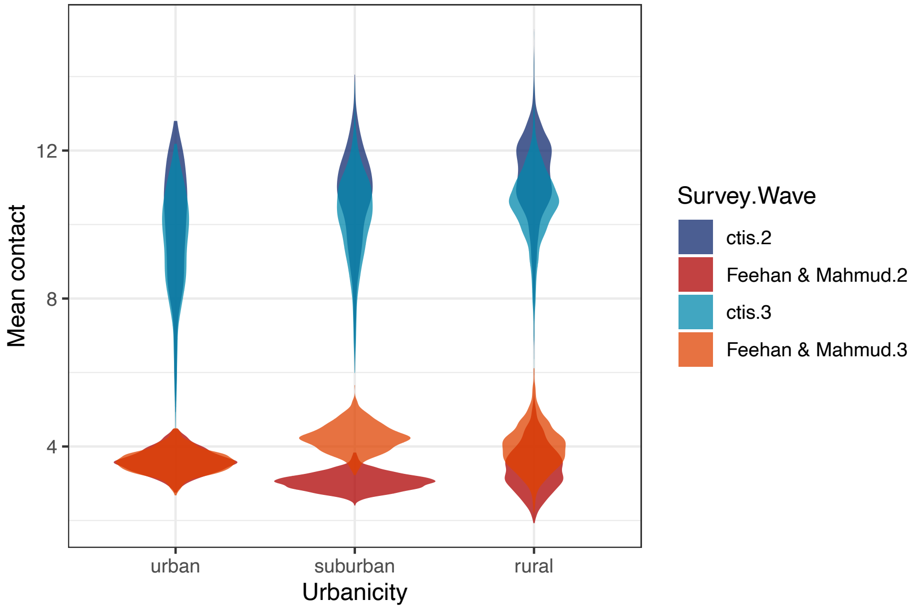


**Figure S14. Comparison between our pandemic contact estimates by urbanicity with those from [27].** Feehan and Mahmud [27] conducted a nationally representative survey of contact patterns in the US including one week in June (Wave 2) and one week in September 2020 (Wave 3). Across the two waves, they had 2,641 respondents who reported fewer than 72 contacts. Respondents were classified as living in urban, suburban, or rural areas. Red and orange violins show the bootstrapped mean contact for June and September 2020, respectively, using provided weights from [27]. (Note that we have imposed a truncation point on their data to match our analysis so the weights may no longer be accurate.) Blue and teal violins show the distribution of mean county estimates from the CTIS for same weeks in June and September, respectively. CTIS data are at the county level; therefore, in this figure counties in NCHS classes 1 and 2 have been designated as urban, 3 and 4 as suburban, and 5 and 6 as rural for visualization purposes.

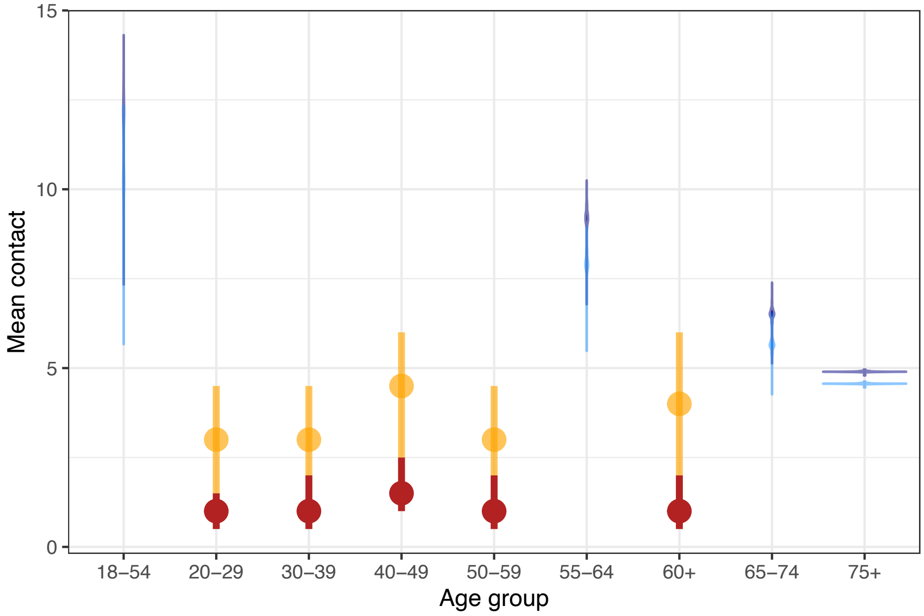


**Figure S15. Comparison between our pandemic contact estimates by age with those from [72].** Kiti and colleagues' surveyed contact patterns across 647 employees of five private companies based in Georgia, USA over two days in April - June 2020 and November 2020 - January 2021. Contacts were categorized as interactions within 6 feet for less than 20 seconds, conversational contacts, or physical contacts, and included contacts in the home. Red and orange point ranges show the means and interquartile ranges of contact divided by two (to represent one day's worth of contacts) for April - June 2020 and November 2020 - January 2021 periods, respectively. Blue and navy violins show the distribution of mean county-age estimates for 158 counties in Georgia from the CTIS for the same time periods, respectively.

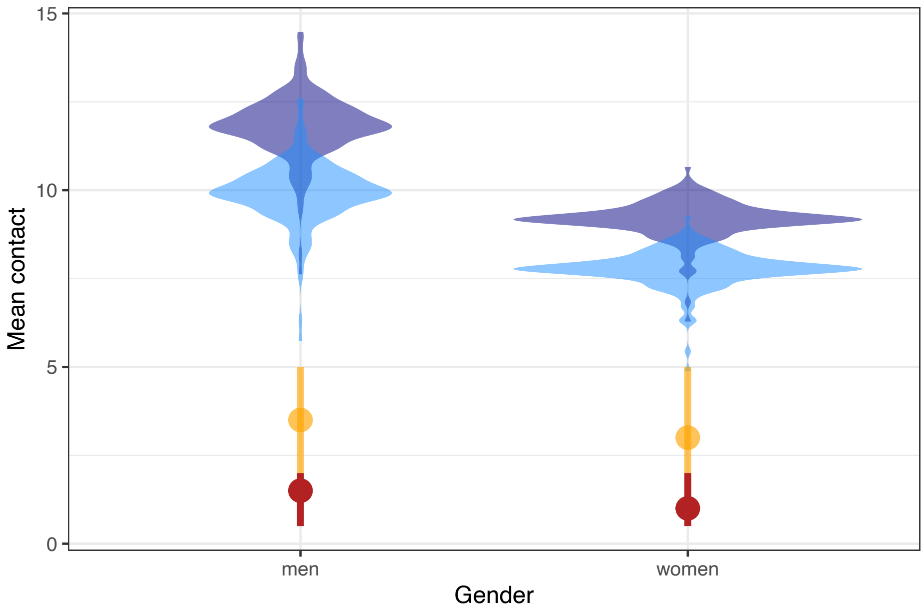


**Figure S16. Comparison between our pandemic contact estimates by gender with those from [72].** Kiti and colleagues' surveyed contact patterns across 647 employees of five private companies based in Georgia, USA over two days in April - June 2020 and November 2020 - January 2021. Contacts were categorized as interactions within 6 feet for less than 20 seconds, conversational contacts, or physical contacts, and included contacts in the home. Red and orange point ranges show the means and interquartile ranges of contact divided by two (to represent one day's worth of contacts) for April - June 2020 and November 2020 - January 2021 periods, respectively. Blue and navy violins show the distribution of mean county-gender estimates for 158 counties in Georgia from the CTIS for the same time periods, respectively.

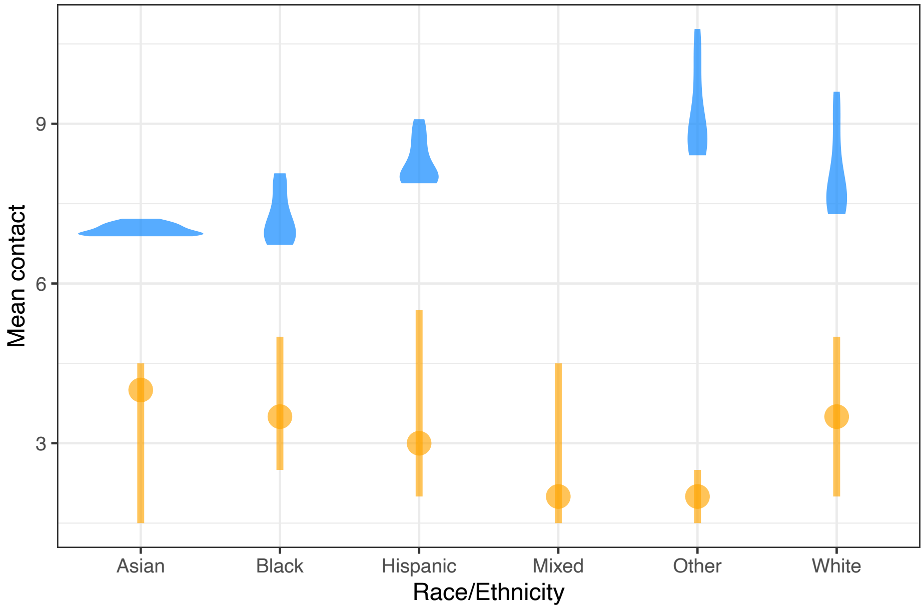


**Figure S17. Comparison between our pandemic contact estimates by race/ethnicity with those from [72].** Kiti and colleagues' surveyed contact patterns across 647 employees of five private companies based in Georgia, USA over two days in November 2020 - January 2021. Contacts were categorized as interactions within 6 feet for less than 20 seconds, conversational contacts, or physical contacts, and included contacts in the home. Orange point ranges show the means and interquartile ranges of contact divided by two (to represent one day's worth of contacts) for November 2020 - January 2021 periods. Blue violins show the distribution of mean state-race/ethnicity estimates for Georgia from the CTIS for each week in the same time period.

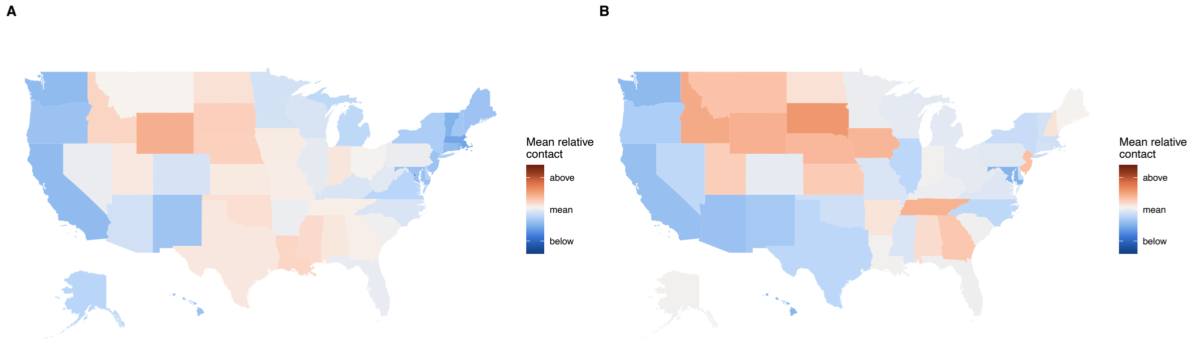


**Figure S18. Comparison between (A) our findings and (B) Breen et al. [22] at the state level.** Relative contact is the mean contact for each state across the study period (May 2020 - May 2021) divided by the national mean across the same period; this normalization allows for a comparison of trends. The contact definition in Breen et al. is the same as [27]; we exclude household contacts for this comparison.

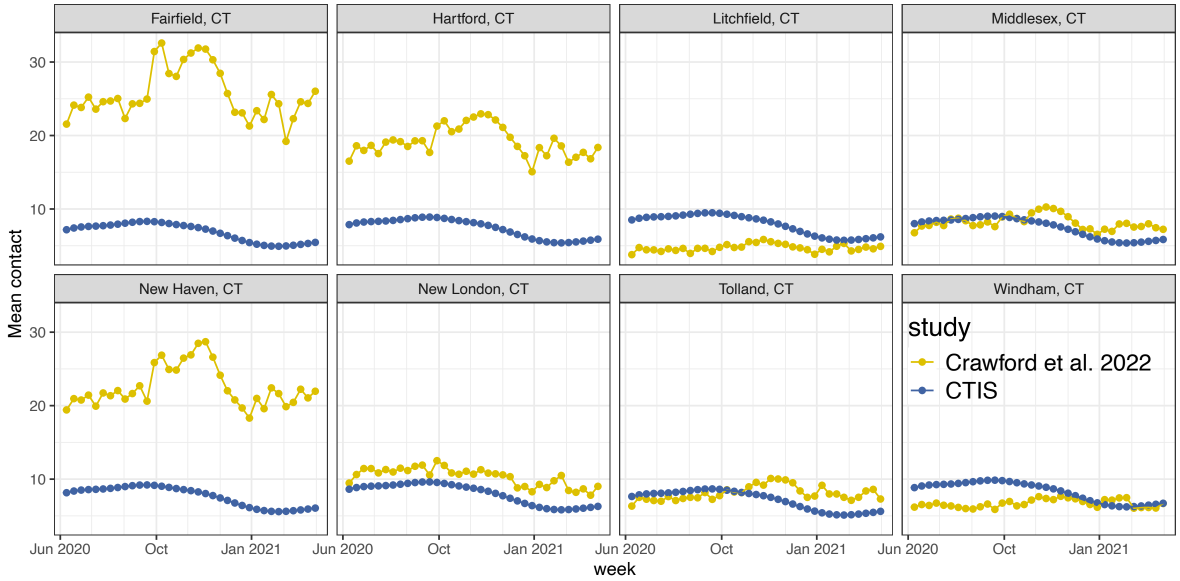


**Figure S19. Comparison between our findings and those from [49].** [49] use mobile device geolocation data in Connecticut to estimate co-location within 2 meters. Here, we aggregate their town level estimates to the county level for comparison using an unweighted mean. [49] yields larger contact estimates than the CTIS, likely due to recall bias in the CTIS, a lower minimum duration for contact in [49], and the potential for double counting contacts with the same person in [49].

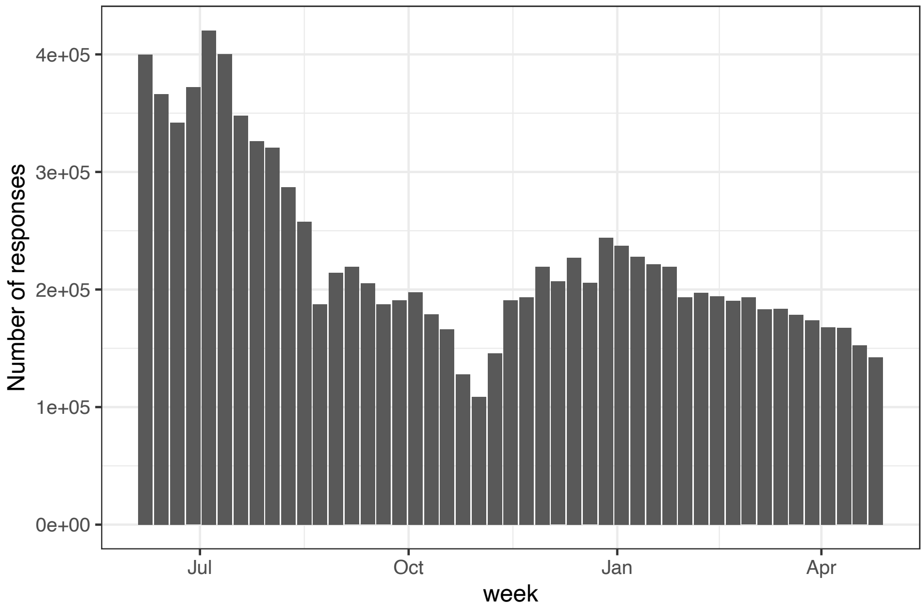


**Figure S20. Weekly valid responses to the CTIS over the study period from June 2020 to April 2021.**

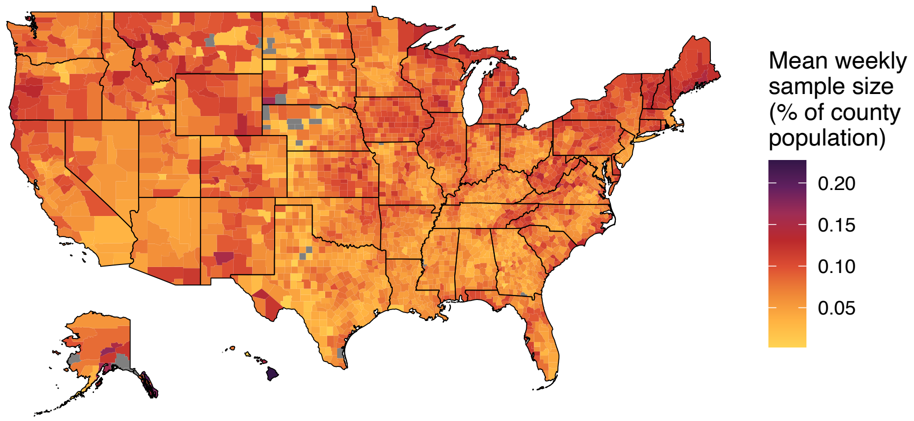


**Figure S21. Mean number of weekly responses per county as a percentage of the county's total population.**

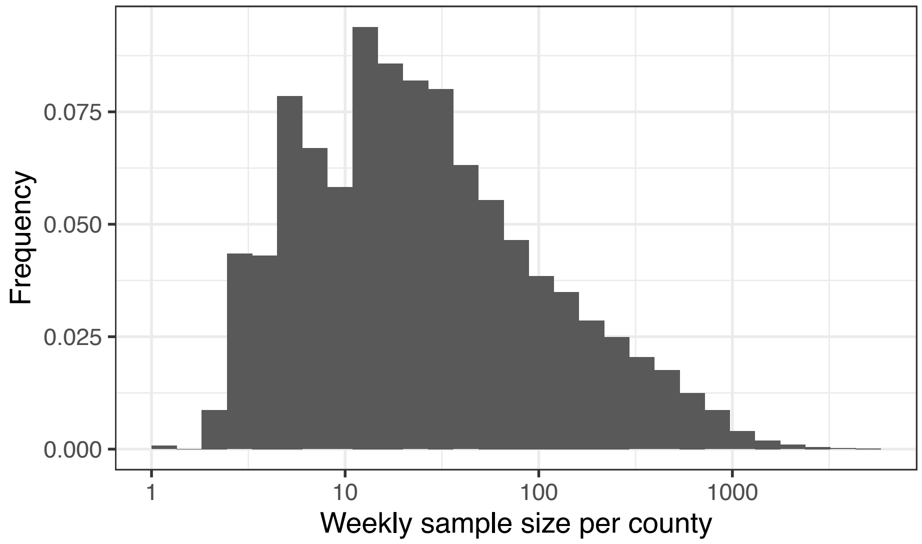


**Figure S22. Weekly sample size by county.** Note that the x-axis is on a log10 scale.

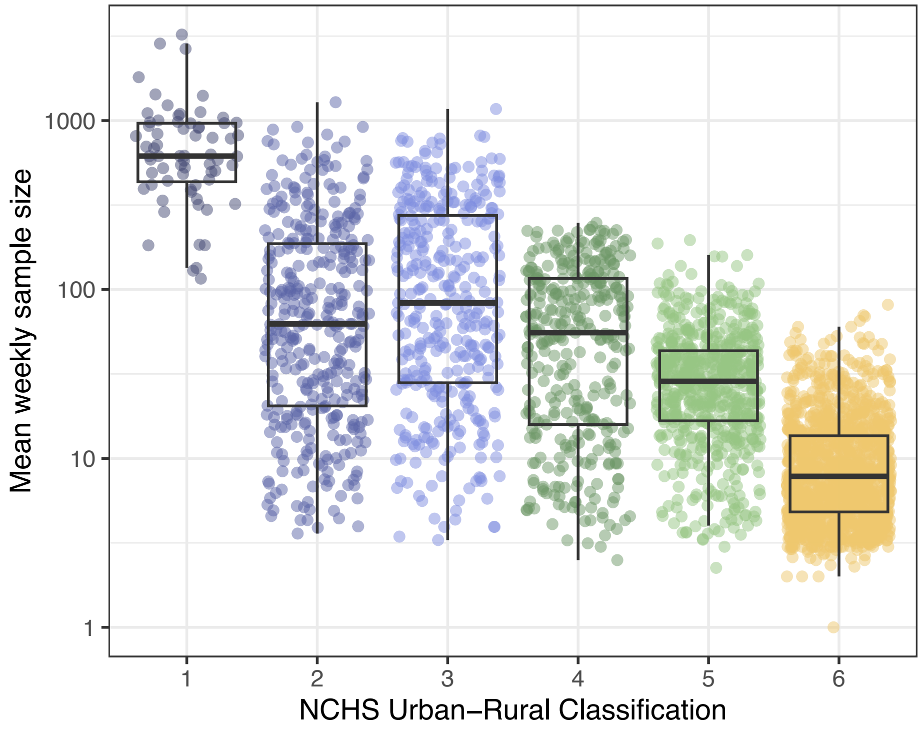


**Figure S23. Weekly sample size by urban-rural designation.** 1 represents the most urban and 6 the most rural. Note that the y-axis is on a log10 scale.

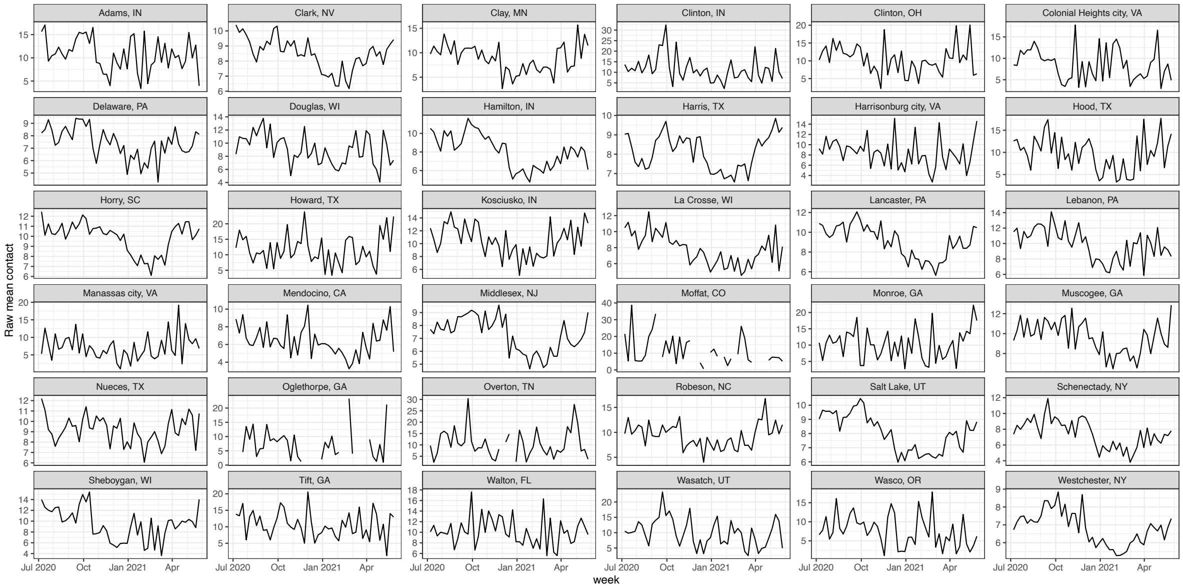


**Figure S24. Example of raw contact time series.** Estimates are very noisy before smoothing but show trend of decreased contact in winter 2020/2021.

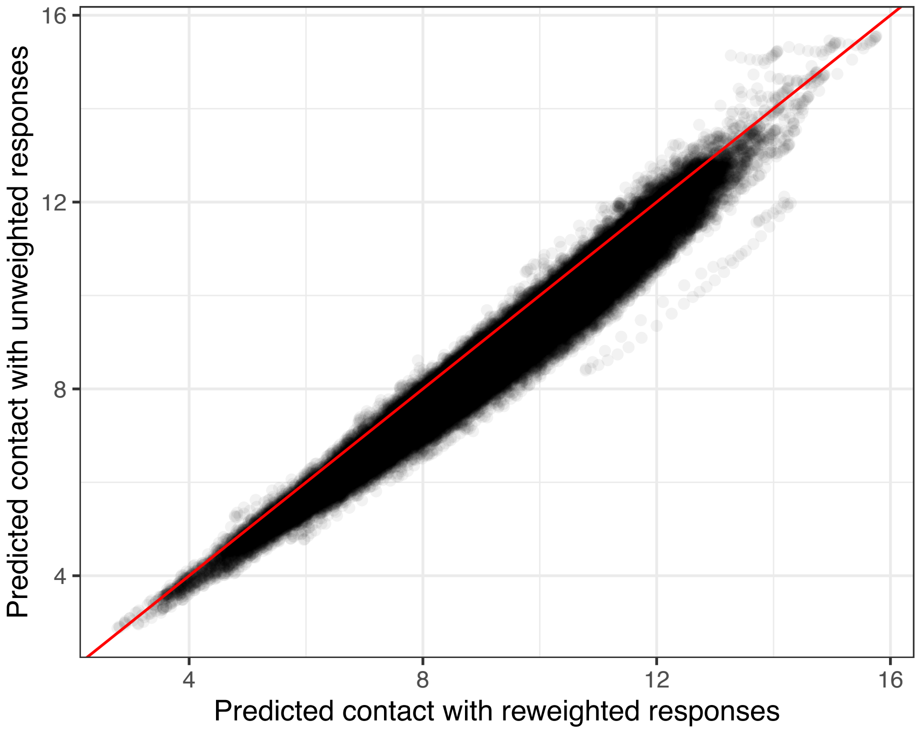


**Figure S25. Comparison of modeled contact from GAMs run on reweighted (post-raking) versus unweighted data.** Contact estimates are slightly higher after reweighting. This finding aligns with our expectation about survey sampling biases: we expect that the survey will disproportionately capture more COVID-19 cautious people with fewer contacts.

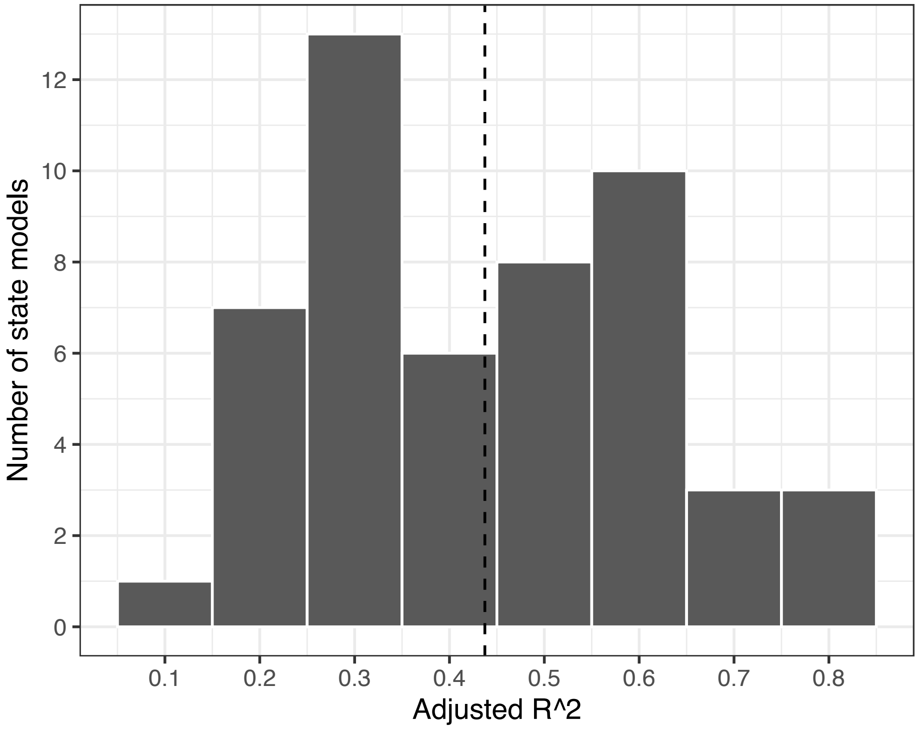


**Figure S26. Distribution of adjusted $R^2$ values across state GAMs.** Dashed line represents the mean adjusted-$R^2$ across states.

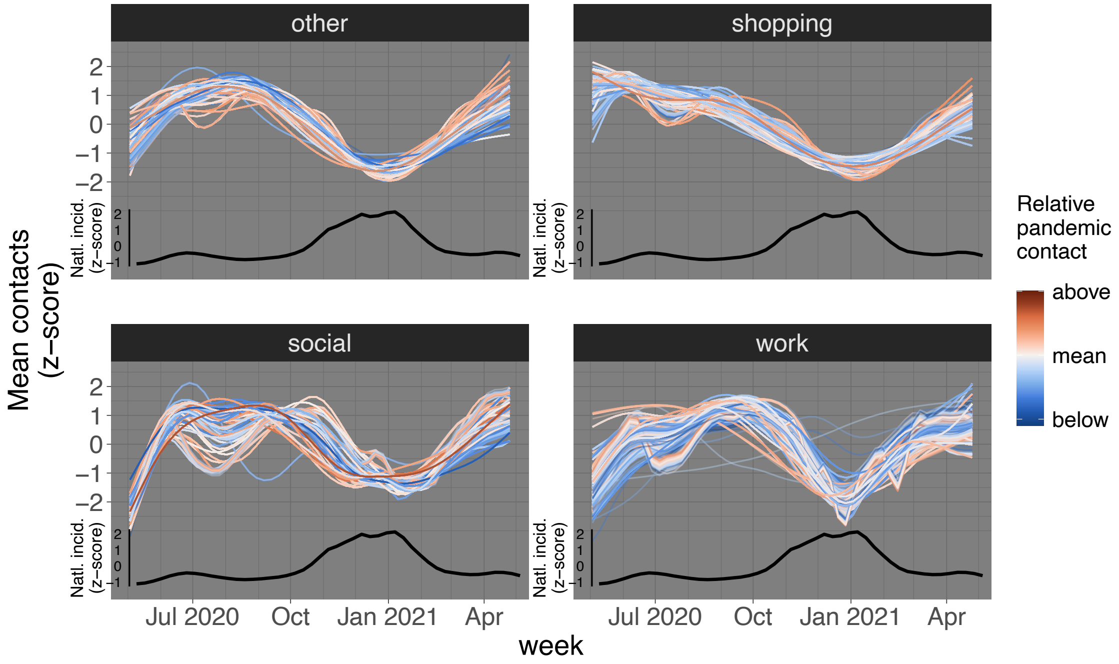


**Figure S27. Time series for contact across settings.** Contact across settings follows the trends of overall contact (summed across settings). Each line represents a county colored by mean contact in that setting relative to the national mean for that setting (above or below); zscored contact relative to each county's mean is shown to allow comparison between time series despite the range of mean contact values across counties. Black line shows the z-score of centered 3-week rolling average of national case incidence for context.

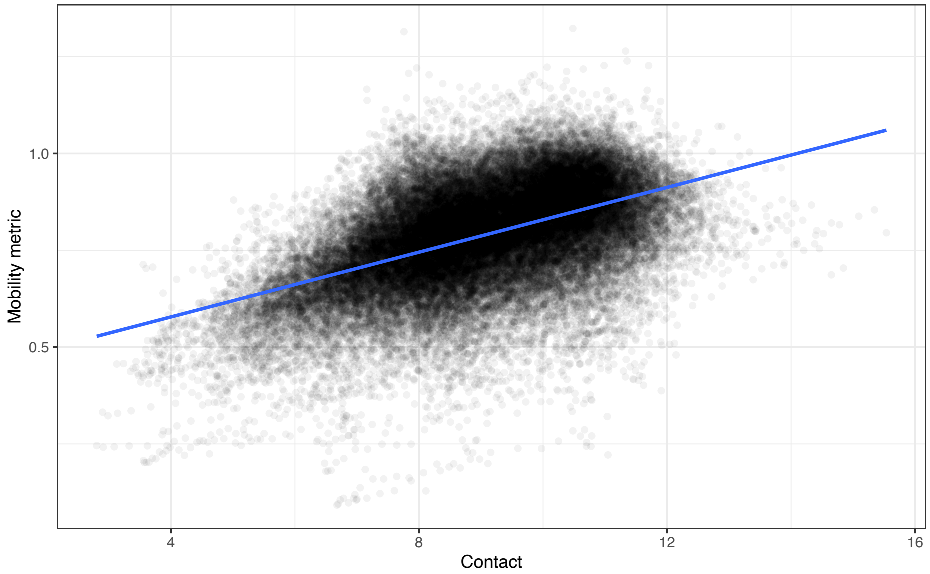


**Figure S28. Mobility and contact are highly correlated.** Data from September - December 2020. Mobility metric is the ratio of weekly 2019 to 2020 Safegraph visitor counts for each county.

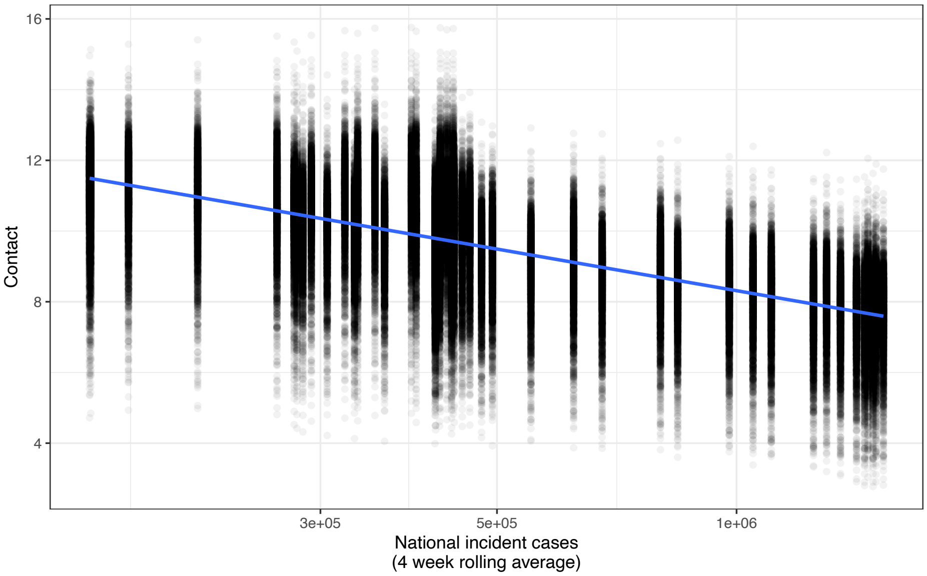


**Figure S29. National incidence and contact are correlated.** Data from June 2020 - April 2021. National incidence is the right-aligned 4 week rolling average.

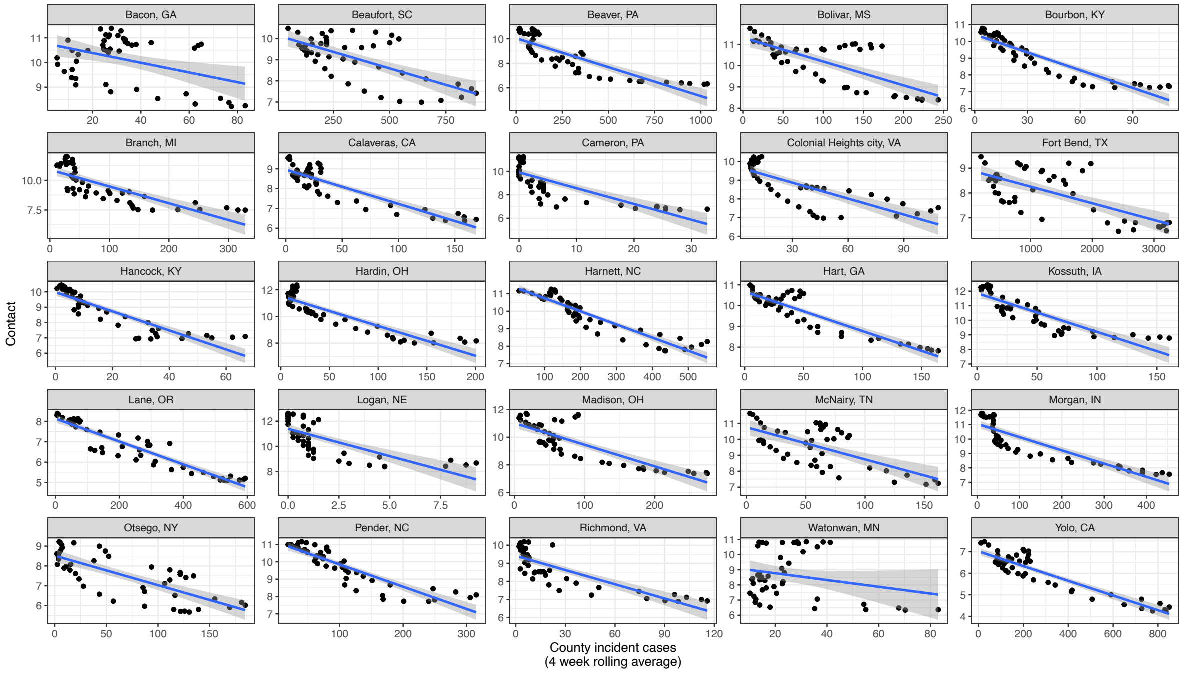


**Figure S30. County incidence and contact are correlated.** Data from June 2020 - April 2021. County incidence is the right-aligned 4 week rolling average.

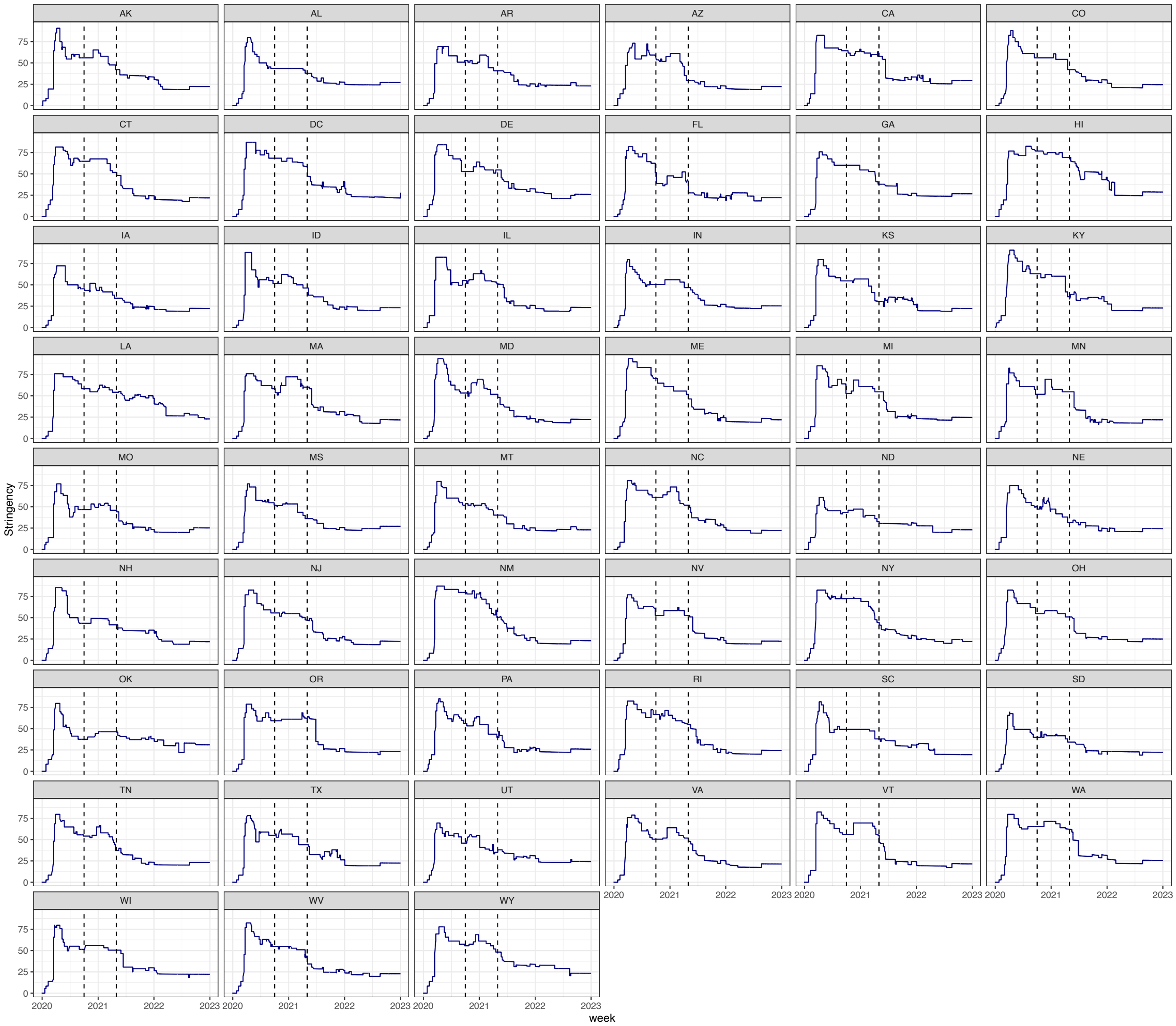


**Figure S31. State stringency indices over time from [40].** Dashed lines denote the beginning and end of our baseline analysis study period.

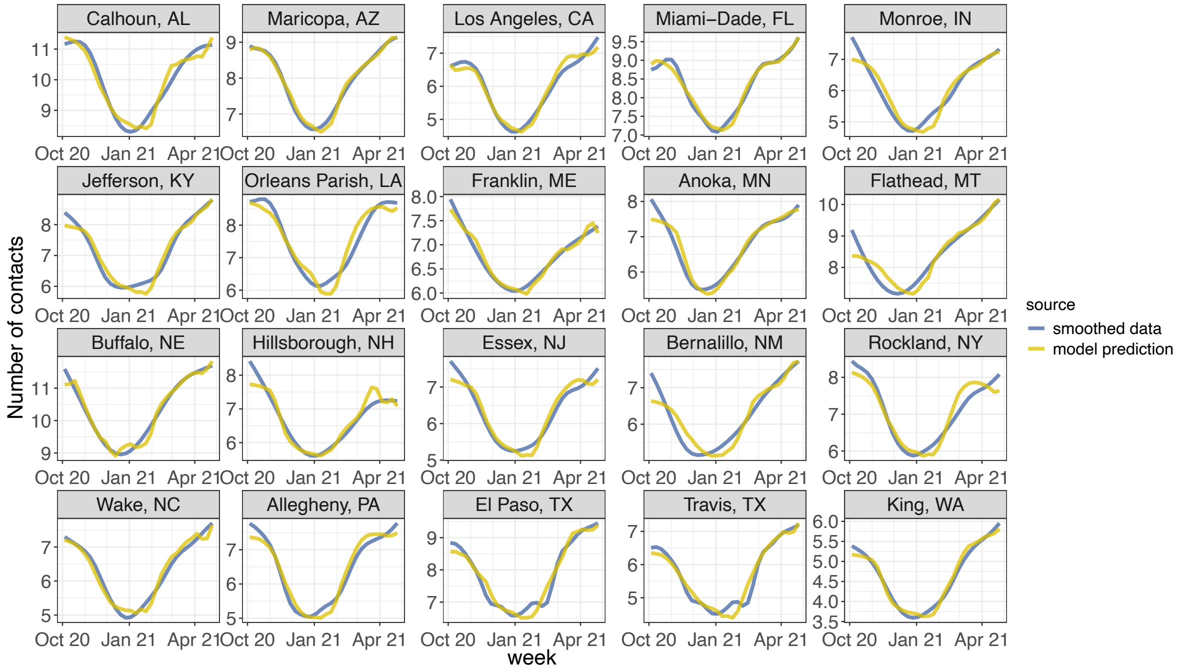


**Figure S32. Smoothed contact data versus linear combination of incidence, policy, and vaccination terms in regression shows good fit between fluctuations in disease and contact.**

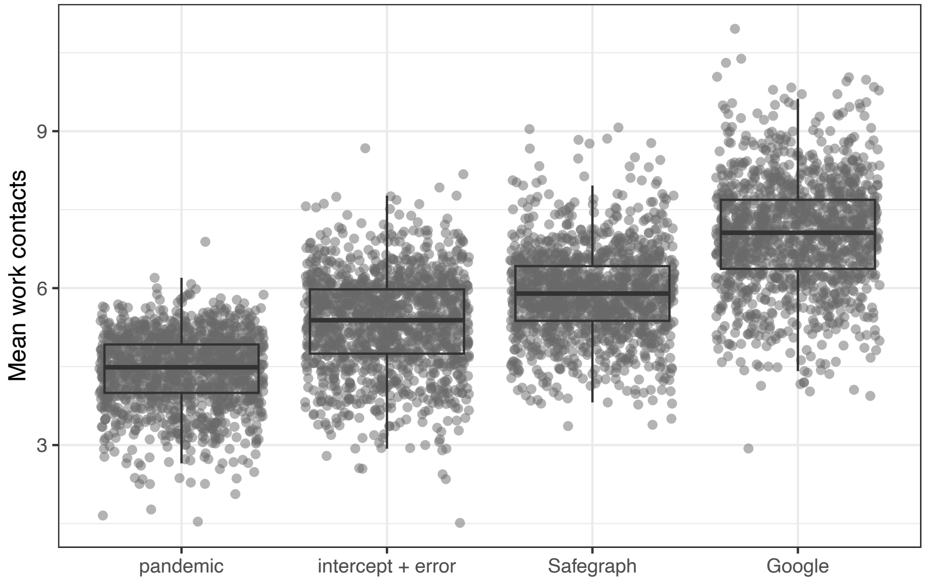


**Figure S33. Using Google work-specific mobility data [76] to infer baseline work contacts yields higher estimates than Safegraph mobility data aggregated across all settings.** Each point represents a county. Intercept + error denotes the intercept from the regression model plus the residual. Safegraph and Google estimates are scaled by changes in mobility data from 2019 to 2020.

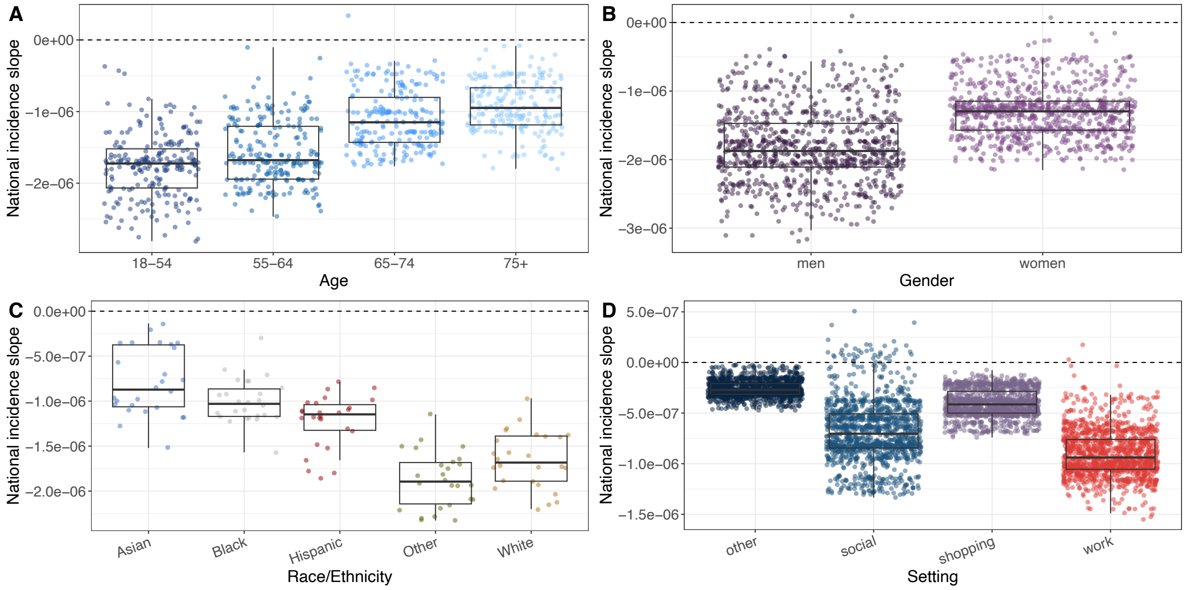


**Figure S34. National disease incidence regression coefficient by age, gender, race/ethnicity, and setting.** More negative coefficients indicate a stronger negative relationship between national incidence and non-household contacts, calculated during the baseline regression.

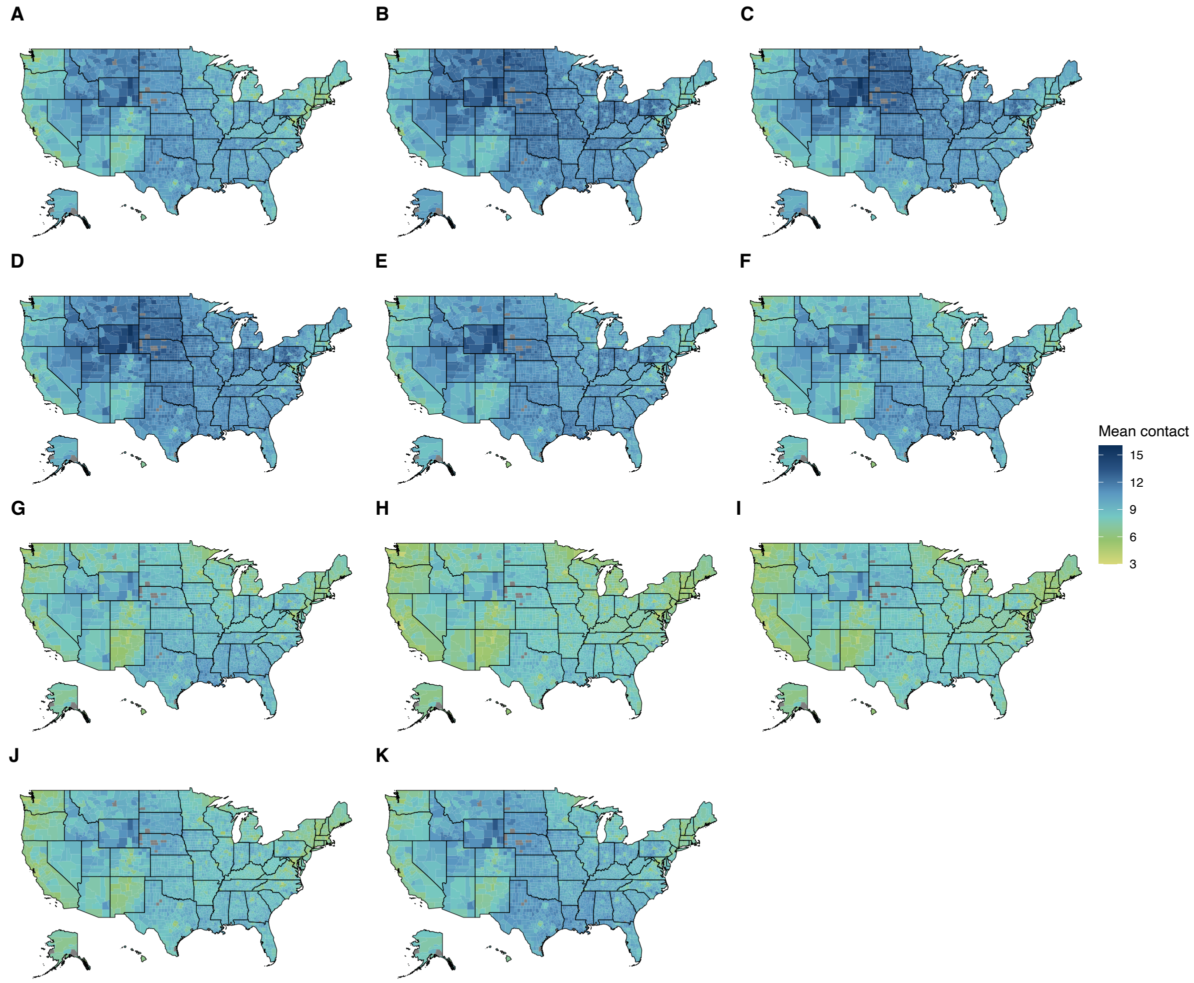


**Figure S35. Fitted contact values by month.** (A) June 2020 through (K) April 2021. While the mean changes, the spatial heterogeneity is relatively consistent.

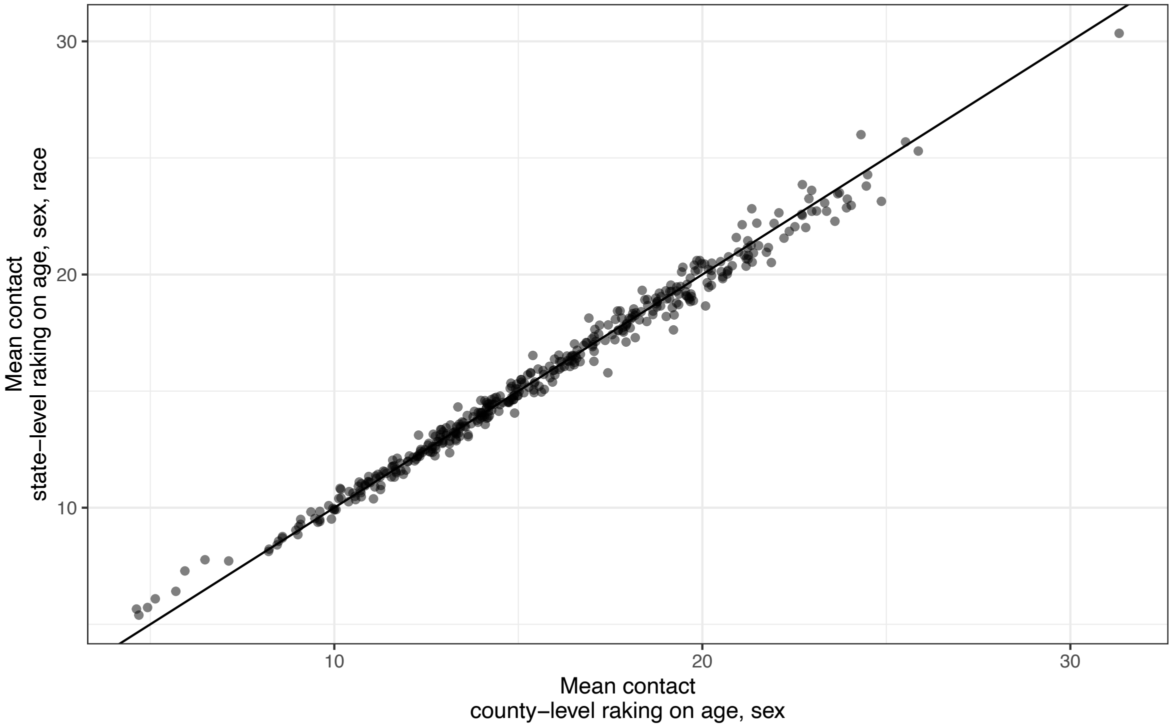


**Figure S36. Raw pandemic contact estimates are minimally affected by raking on race in addition to age and gender.** X-axis value is generated by raking at the county-level on age and gender and aggregating estimates to the state-month level. Y-axis value is generated by raking at the state-level on age, gender, and race and aggregating estimates to the monthly level.

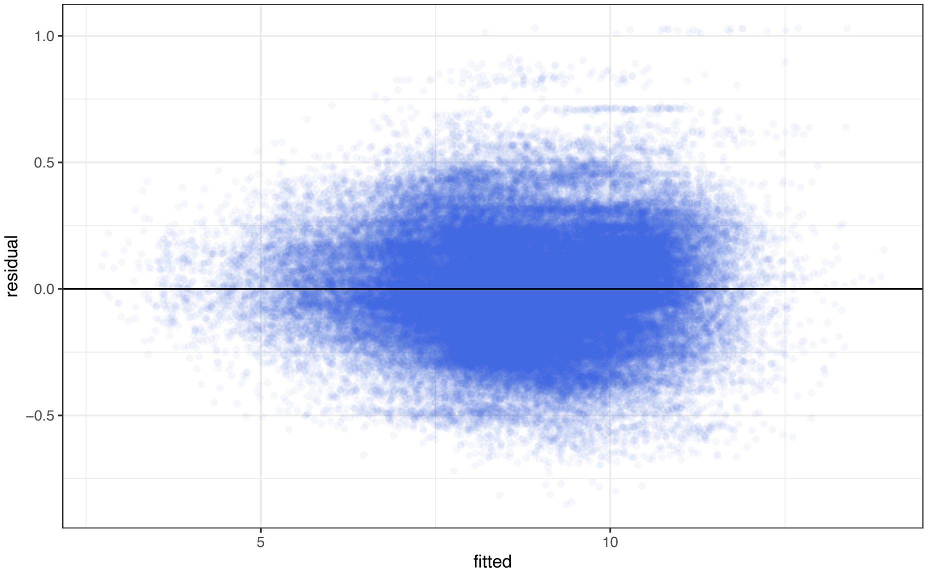


**Figure S37. Residuals versus fitted values for baseline regression across all counties indicate no clear trends nor fanning/heteroscedascity.**

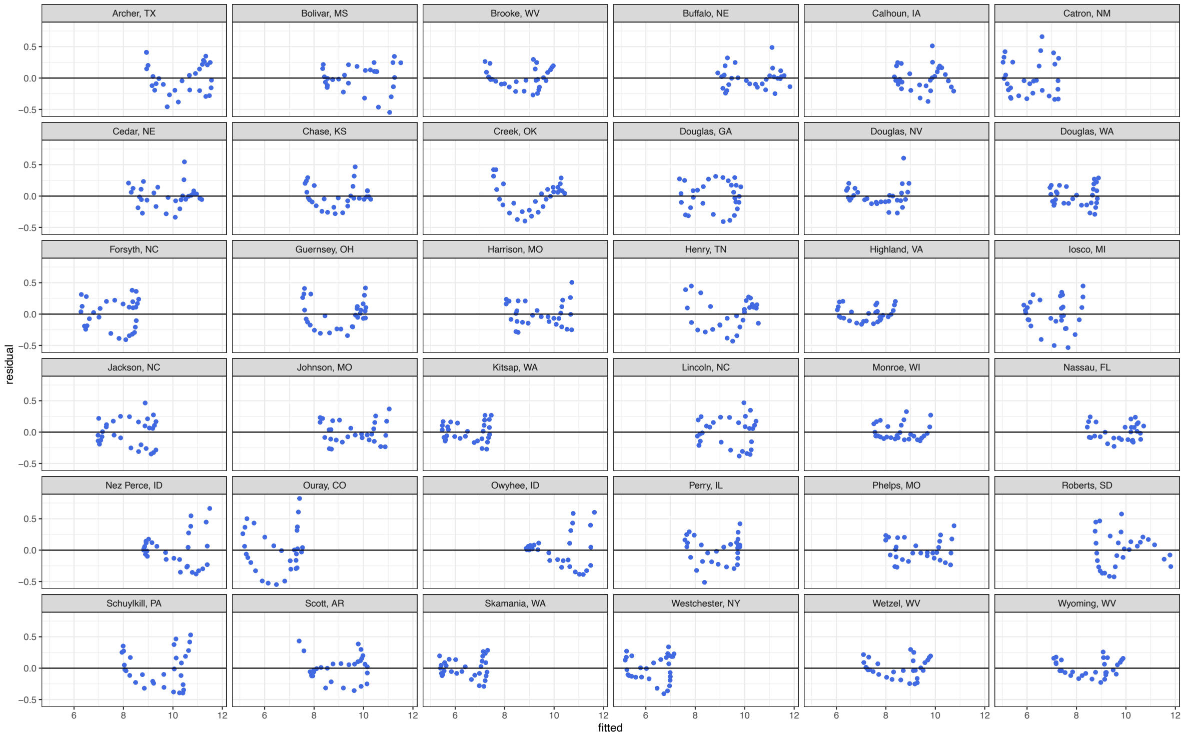


**Figure S38. Residuals versus fitted values for a sample of random counties.**

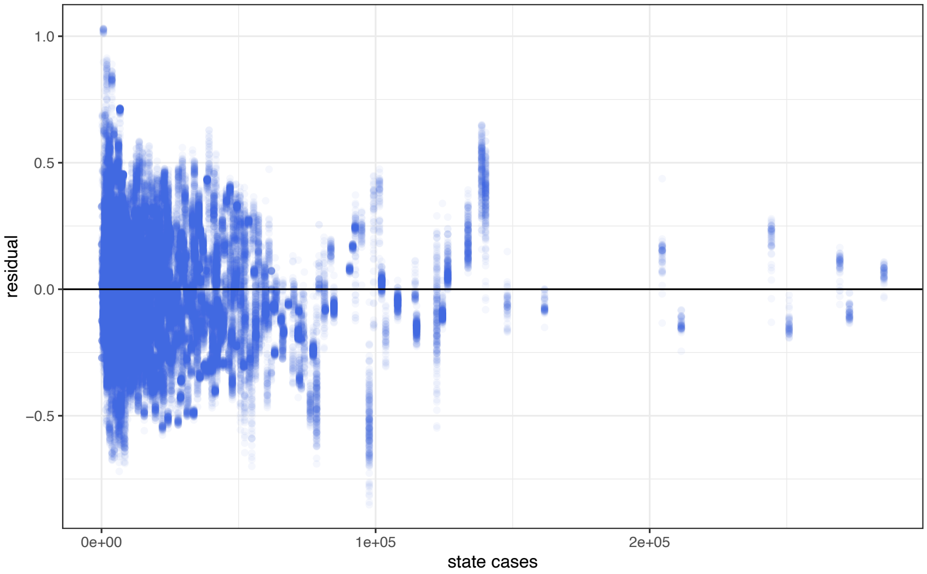


**Figure S39. Residuals from baseline regression models versus state case incidence demonstrates no clear relationship, suggesting that it is reasonable to exclude state cases from the models.**

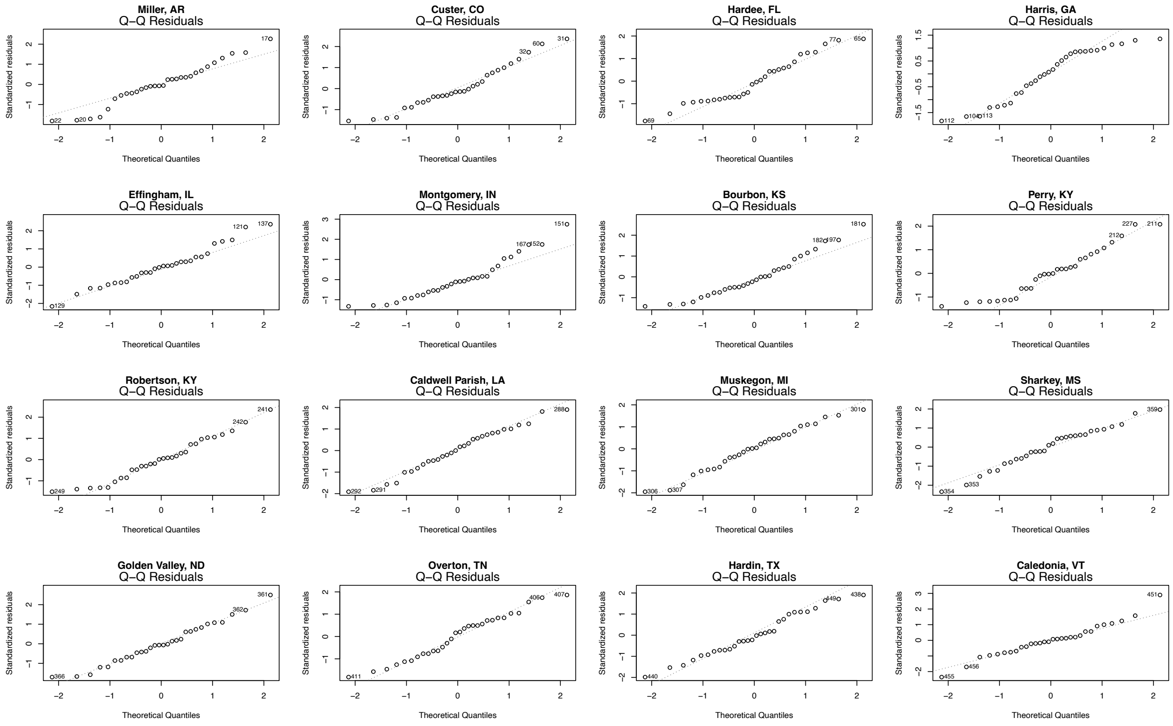


**Figure S40. QQ-plots for a random sample of counties demonstrate that residuals are plausibly normal.**

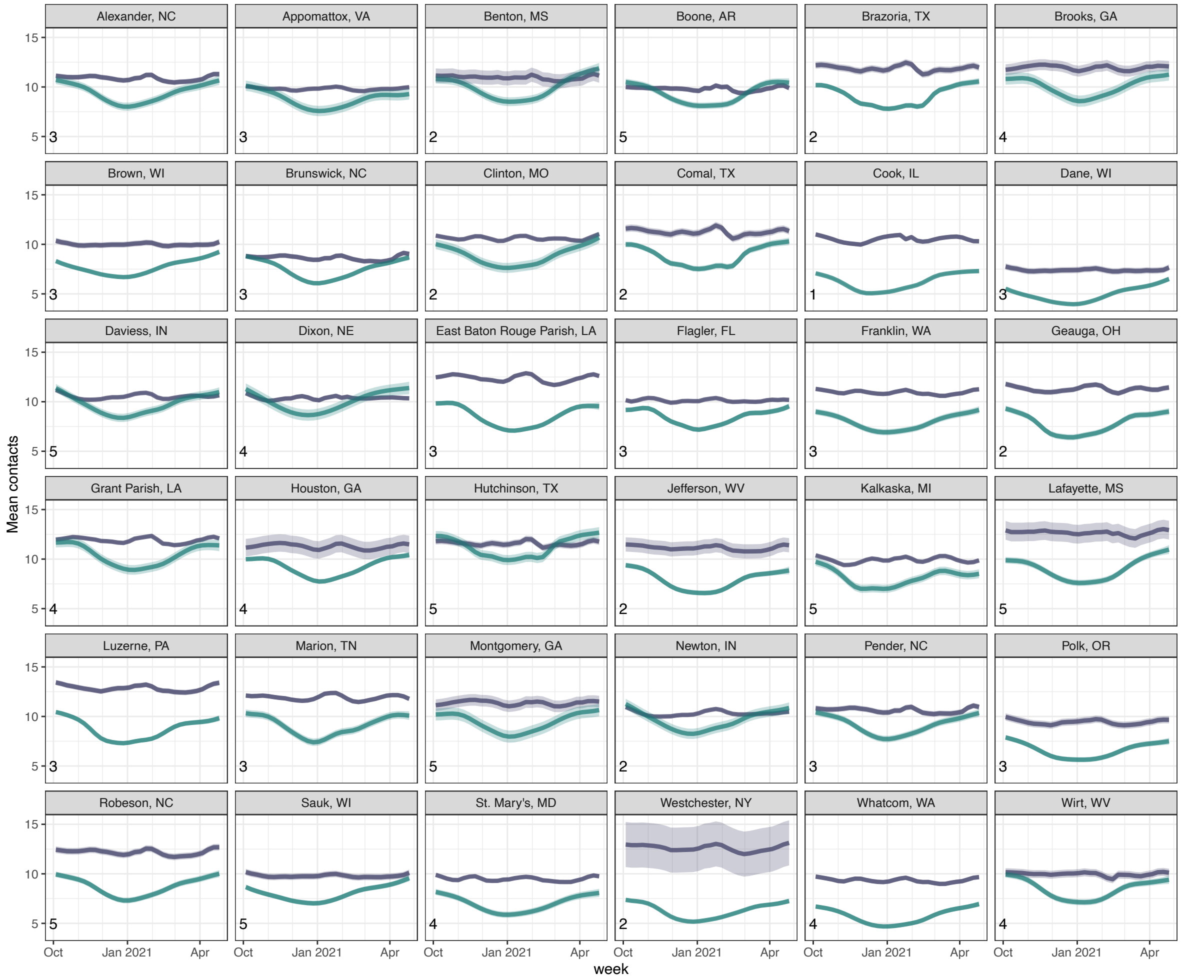


**Figure S41. Non-pandemic vs pandemic trajectories for a random sample of counties spanning different urbanicity levels.** NCHS classification shown in lower left corner where 1 is urban and 6 rural. Non-pandemic predictions are shown in slate and pandemic estimates are in teal.

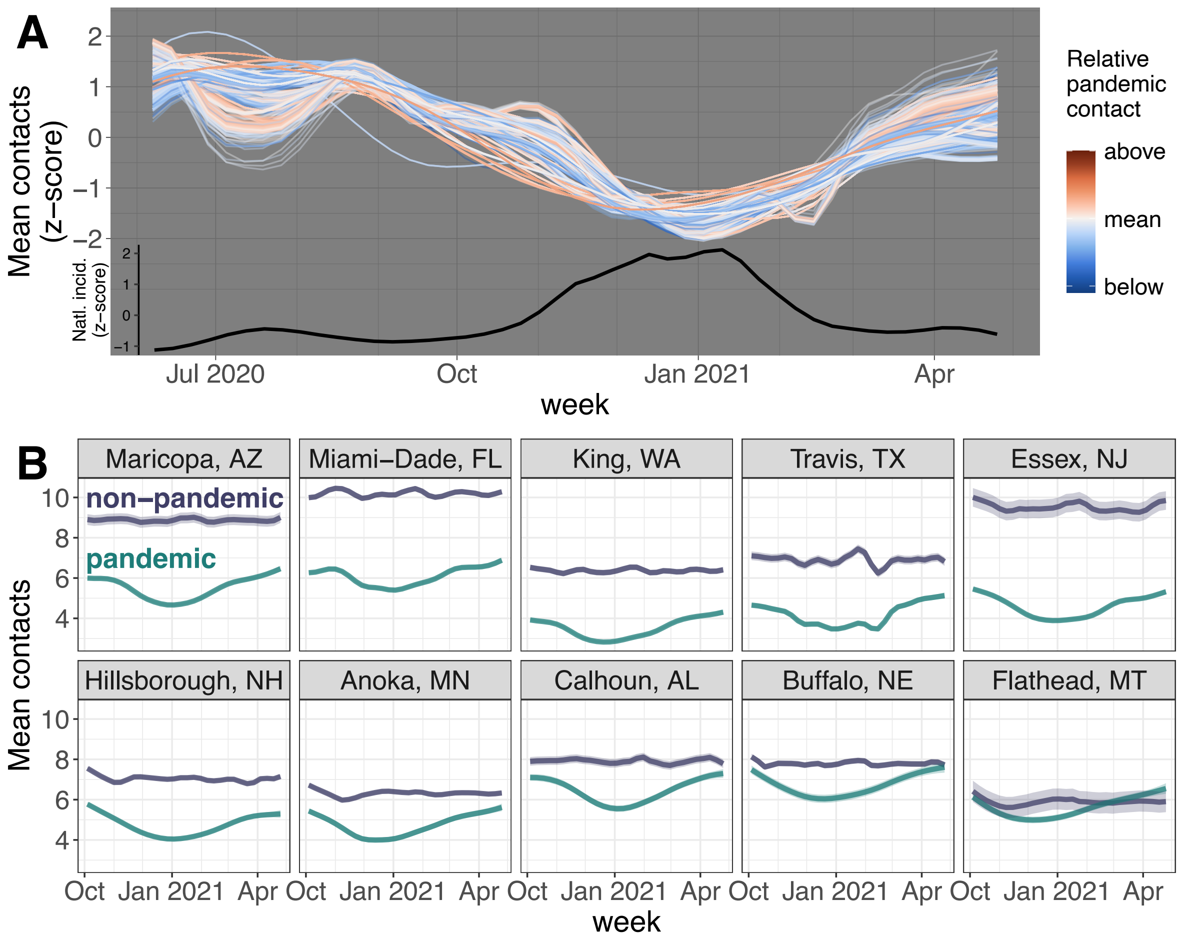


**Figure S42. Counties have similar contact dynamics over time and relatively stable contact after controlling for disease using 90th percentile truncation (36 contacts).** For more figure details, compare to and see caption for Figure 1 in the main text.

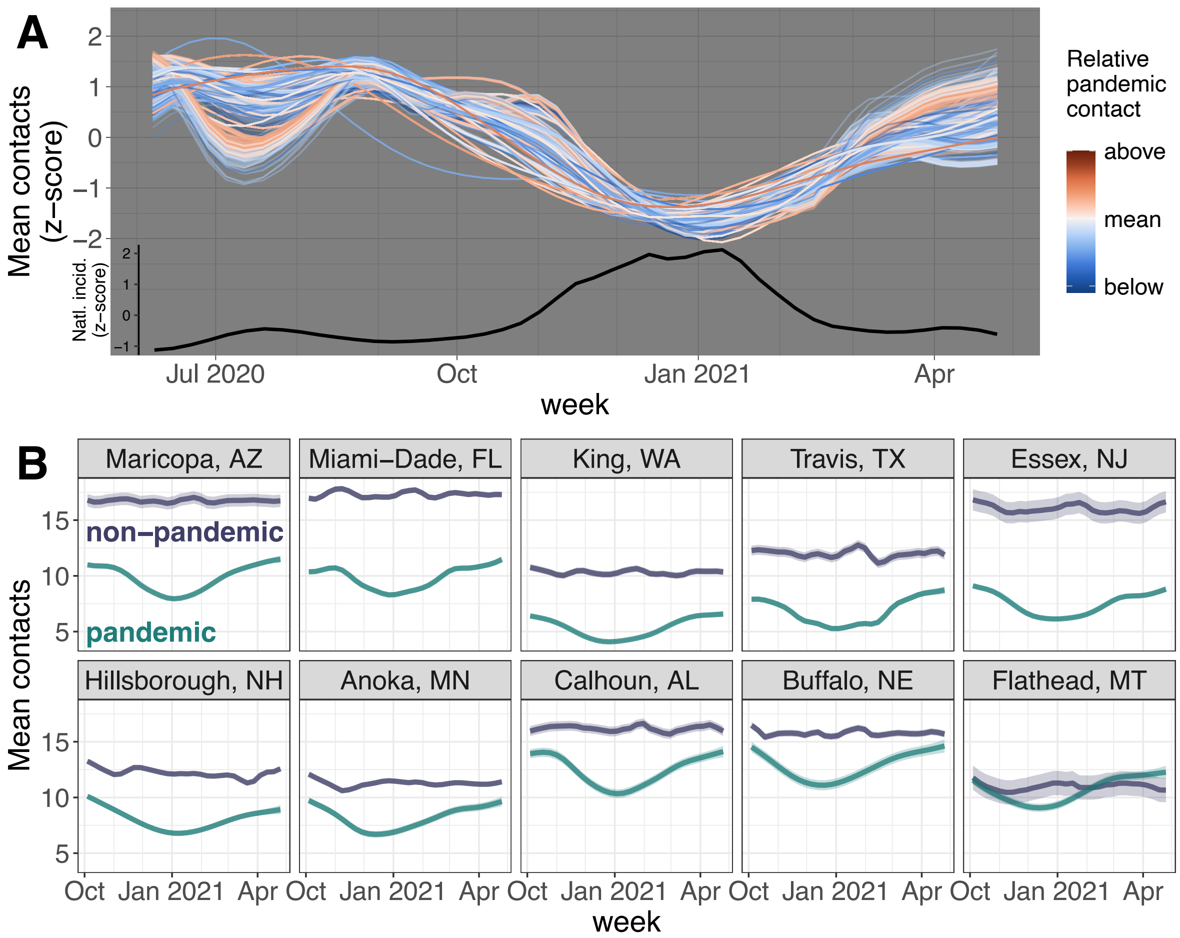


**Figure S43. Counties have similar contact dynamics over time and relatively stable contact after controlling for disease using 97th percentile truncation (108 contacts).** For more figure details, compare to and see caption for Figure 1 in the main text.

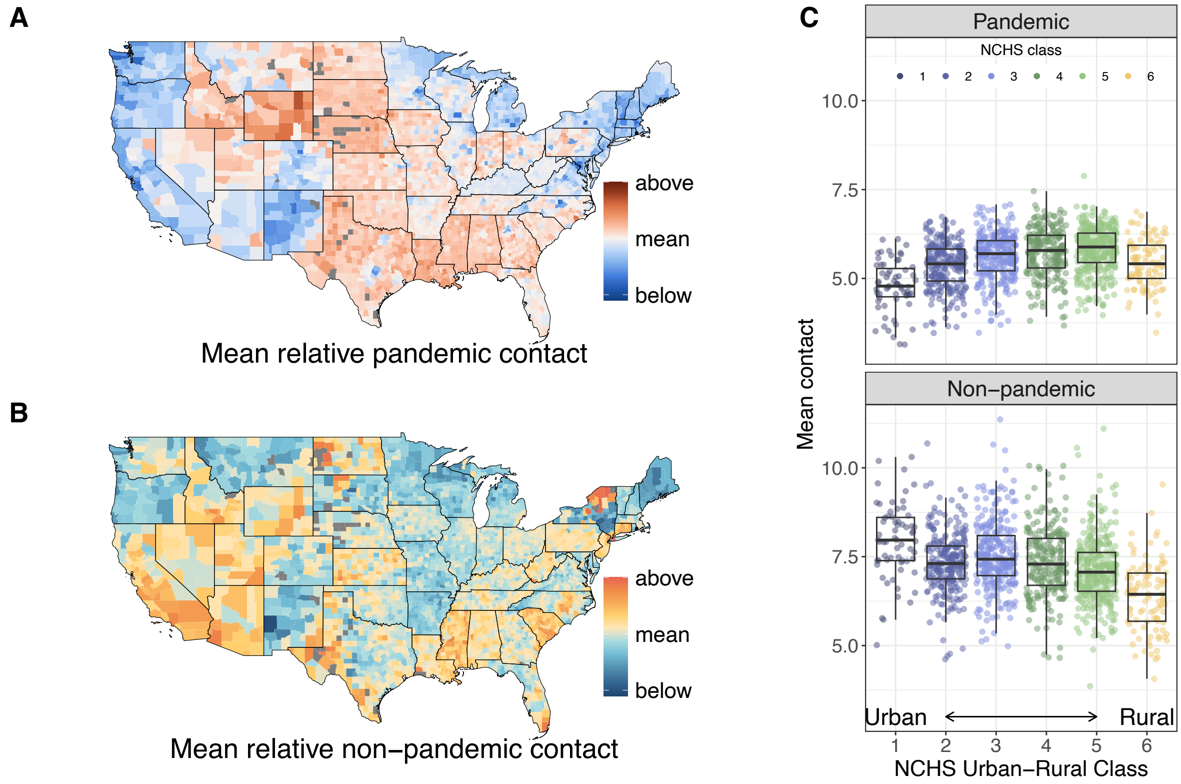


**Figure S44. Contact is spatially heterogeneous regardless of disease incidence, but the urban-rural gradient reverses after controlling for disease incidence using 90th percentile truncation point (36 contacts).** Reversal of pandemic urban-rural gradient is stronger with this truncation than with the 95th percentile in the main text. For more figure details, compare to and see caption for Figure 2 in the main text.

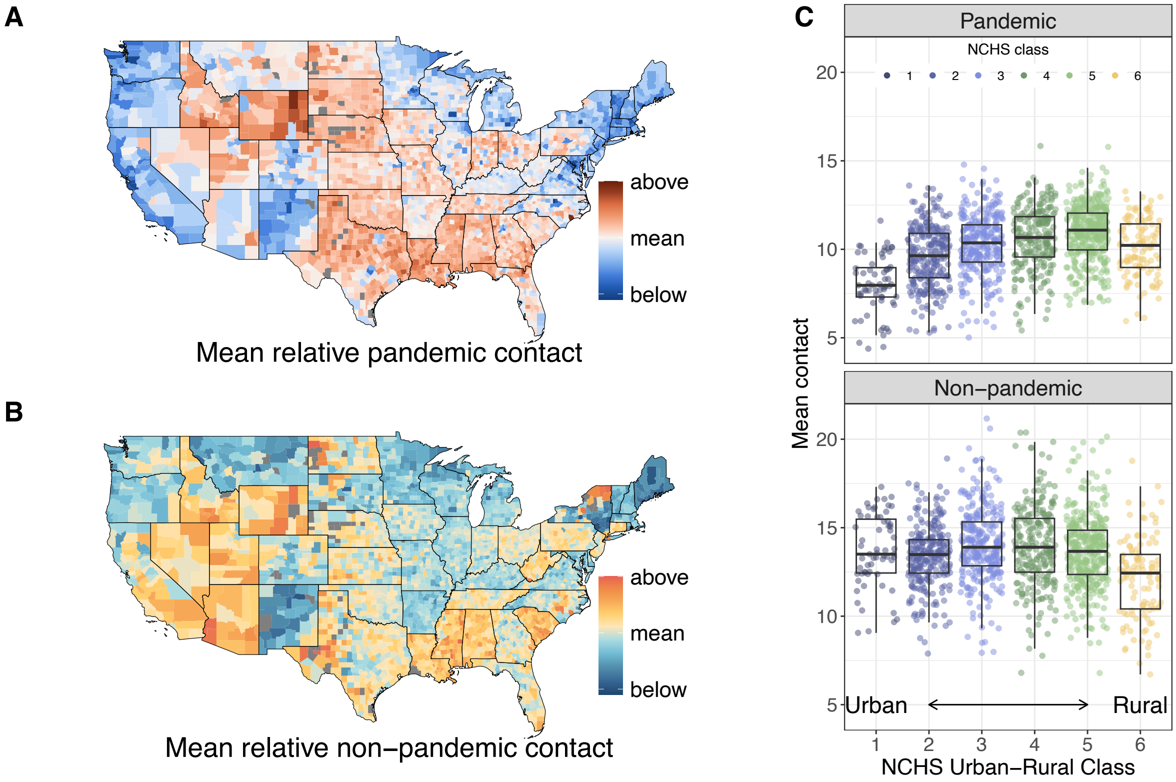


**Figure S45. Contact is spatially heterogeneous regardless of disease incidence, but the urban-rural gradient reverses after controlling for disease incidence using 97th percentile truncation point (108 contacts).** For more figure details, compare to and see caption for Figure 2 in the main text.

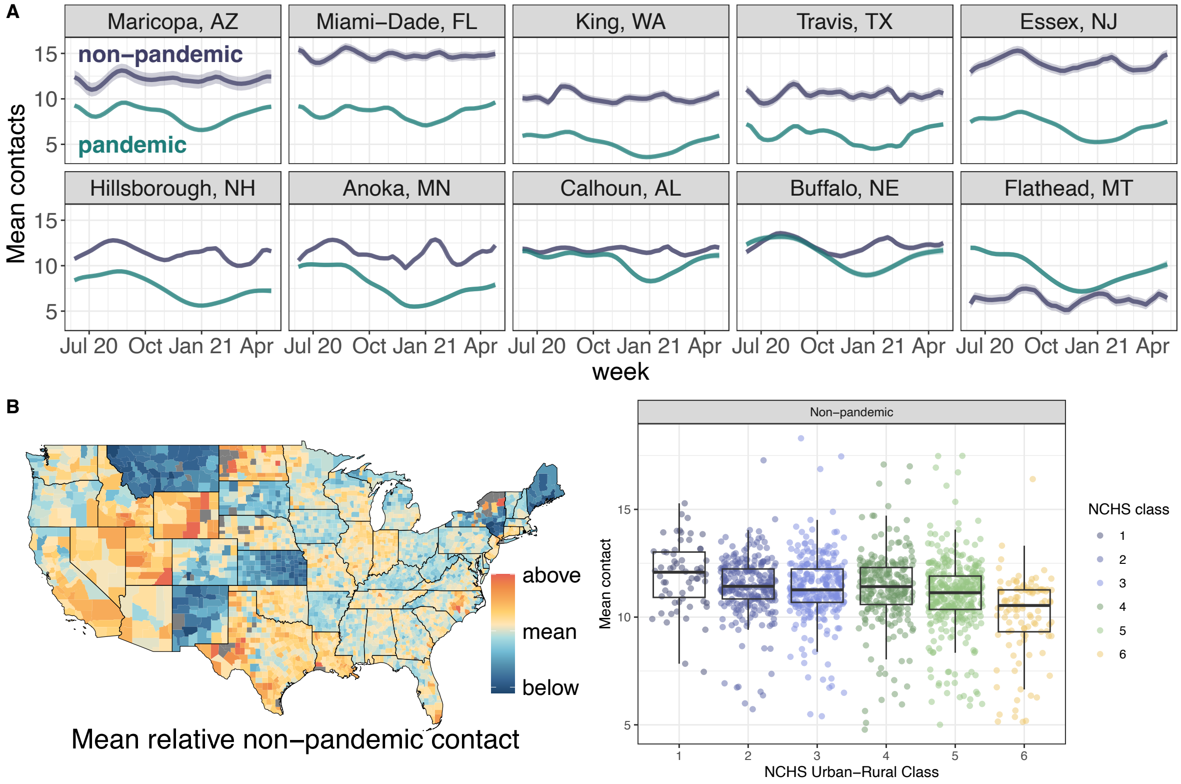


**Figure S46. Baseline estimates are less stable when contact data are fit over a longer period (June 2020 - April 2021) with more variation in incidence and policy levels.**

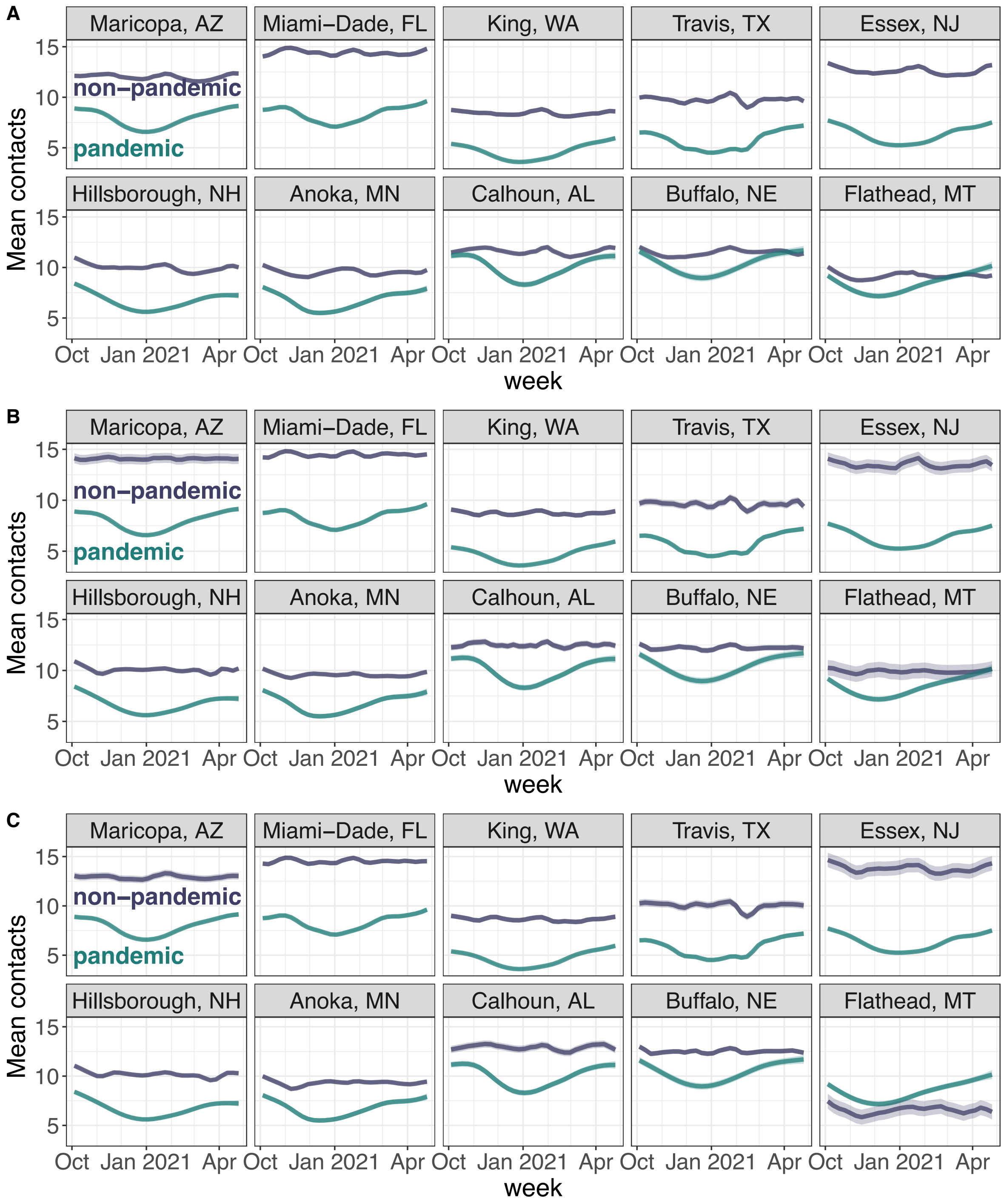


**Figure S47. Estimates of baseline contact over time are consistent across different regression models.** (A) Model using solely national incident cases and county vaccination coverage, i.e. no policy data. (B) Model incorporating county-level incident cases in addition to covariates in main text model. (C) Same model as main text but without interaction between national incident cases and county vaccination coverage.

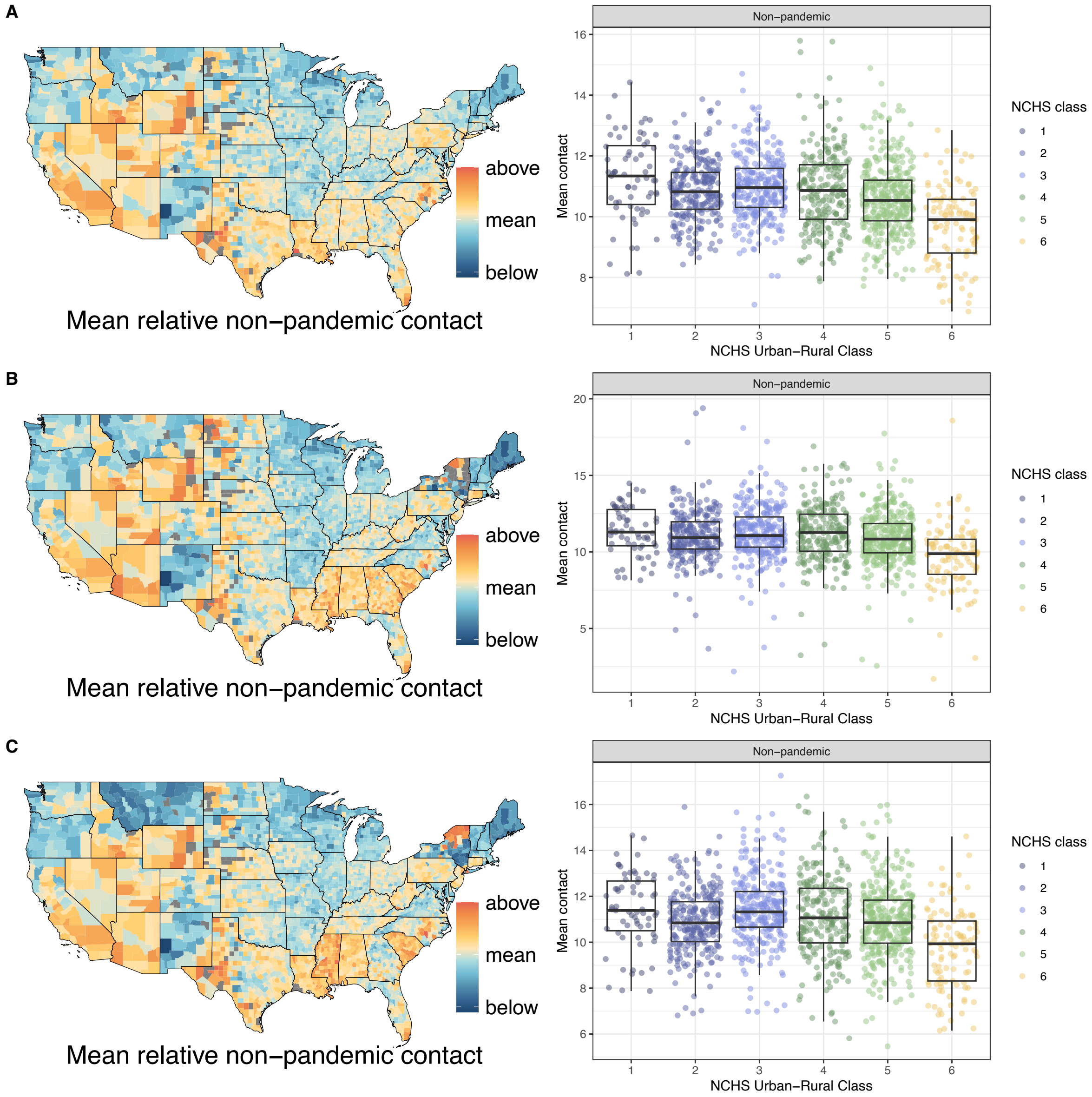


**Figure S48. Estimates of baseline contact across space and urbanicity are consistent across different regression models.** (A) Model using solely national incident cases and county vaccination coverage, i.e. no policy data. (B) Model incorporating county-level incident cases in addition to covariates in main text model. (C) Same model as main text but without interaction between national incident cases and county vaccination coverage.

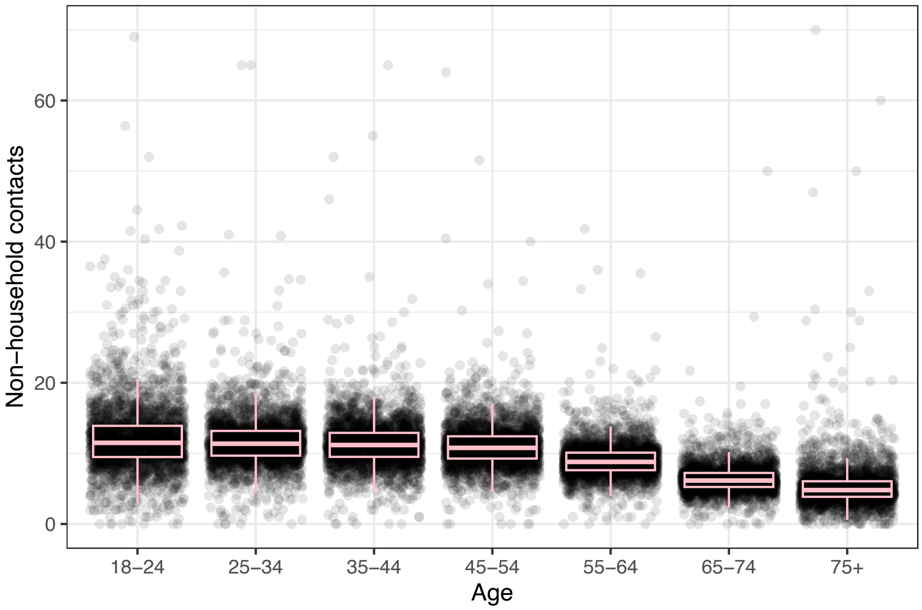


**Figure S49. Age groups 18-54 have roughly the same number of contacts.** Each point represents a county, raw mean contact for the period June 2020 through April 2021.

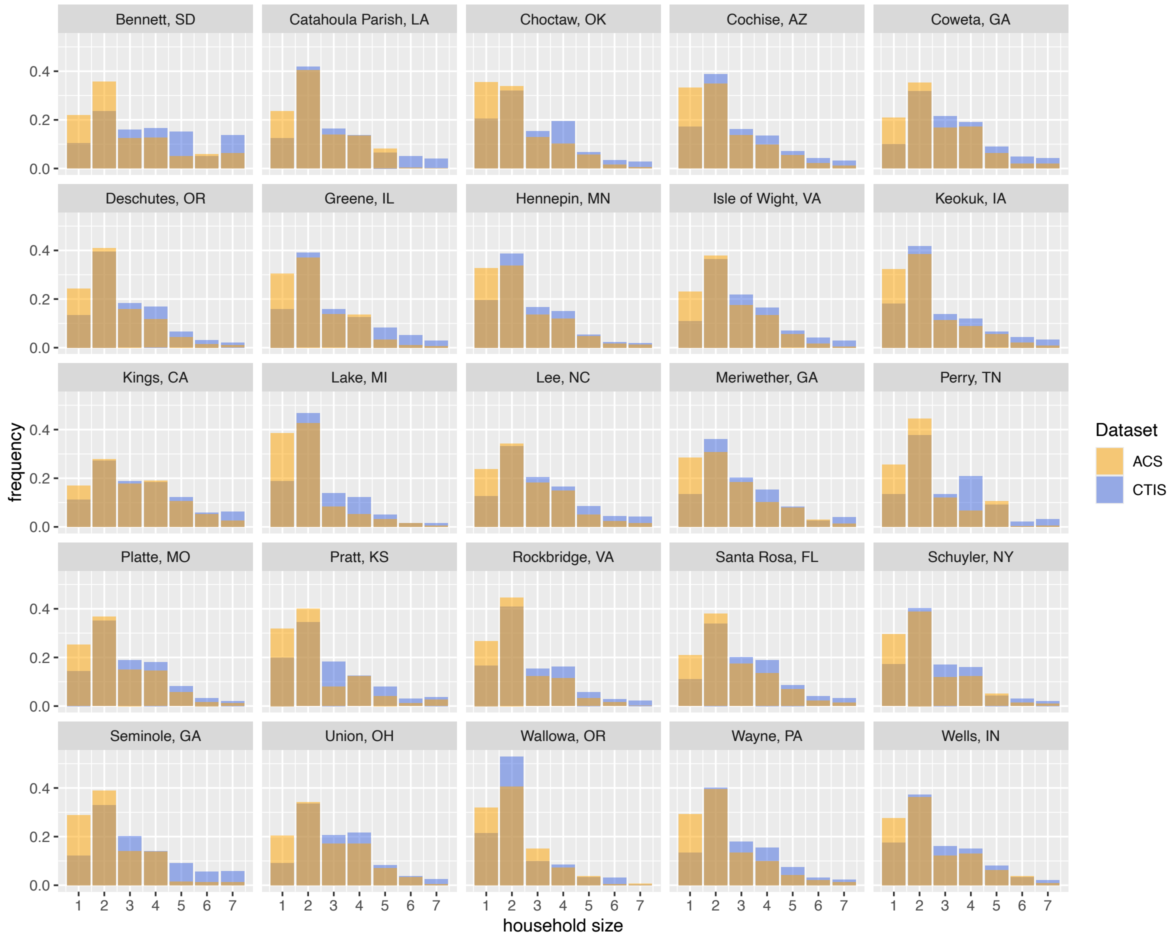


**Figure S50. Comparison between household size in raked contact data and census data for a subsample of counties shows that the CTIS decently captures range of household sizes and we do not need to rake on household size.** In general, smaller households are underrepresented in the survey and larger households are overrepresented, as expected given that larger households have more people that can respond to the survey.

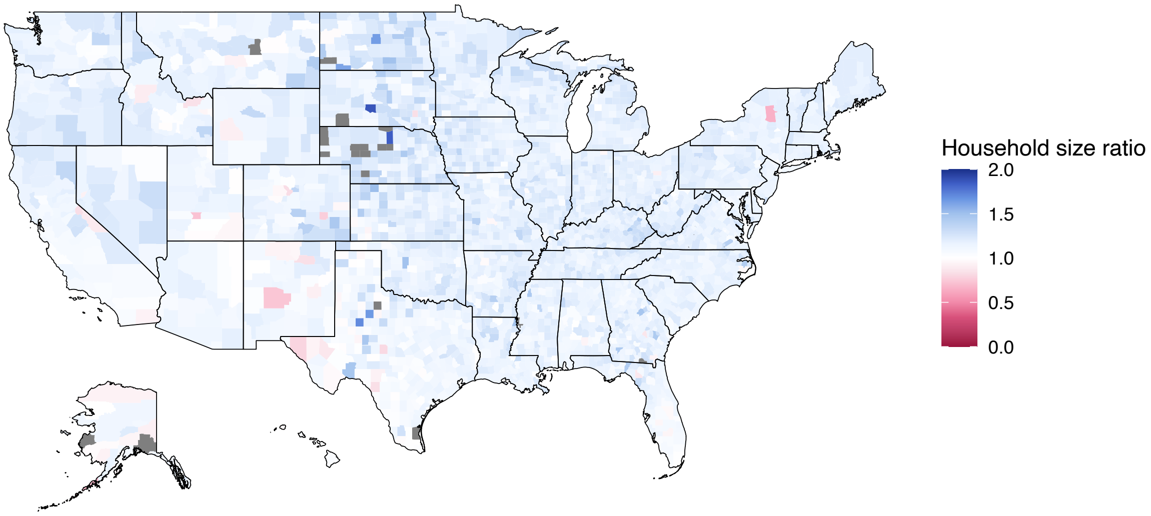


**Figure S51. Ratios of mean survey household size and census household size are close to 1.** This suggests we do not need to rake on household size as our sample is fairly representative.

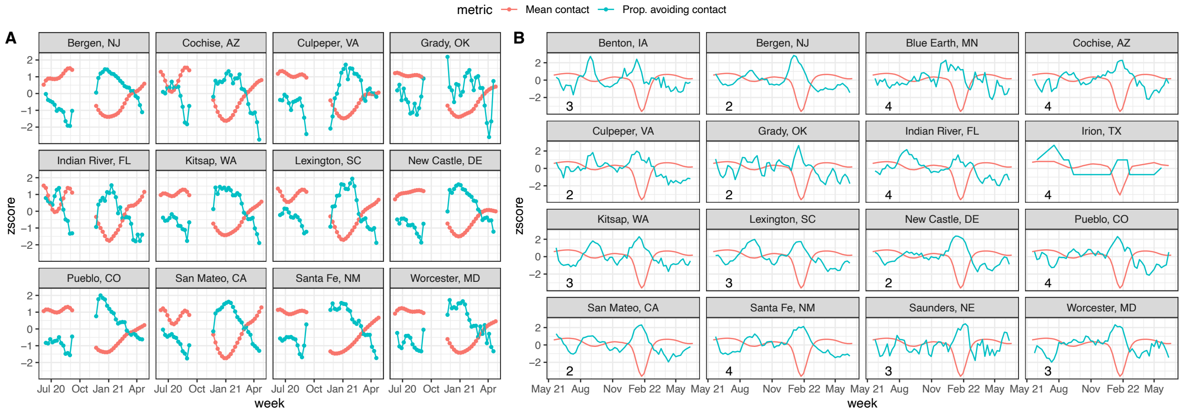


**Figure S52. Contact is inversely correlated with the proportion of individuals avoiding contact, as reported in the CTIS. Predicted contact based on national incidence maintains this relationship with observed avoidance data.** (A) Z-scored centered 3-week rolling average of raw proportions of respondents avoiding contact compared to z-scored GAM-estimated contact. The question about avoiding contacts was omitted from the CTIS from Step 8, 2020 to November 24, 2020, hence the gap in data. (B) Predicted contact (based only on national case incidence) compared to reported proportion avoiding contact on the CTIS from May 2021 through the end of the survey in June 2022.

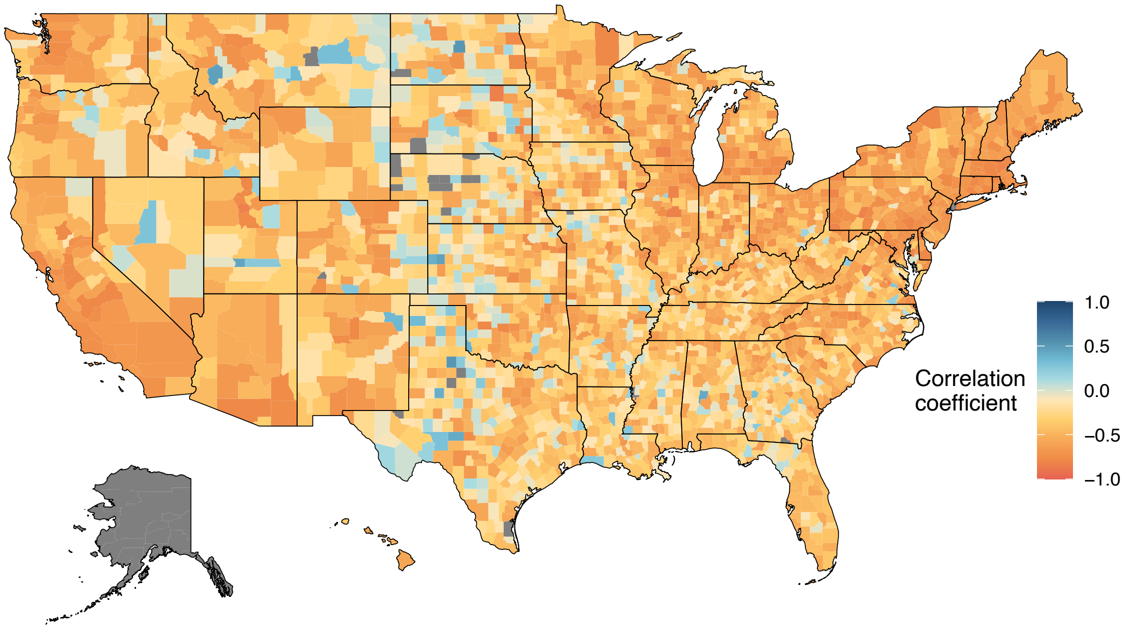


**Figure S53. Correlation between predicted out of sample contact and observed contact avoidance across counties.** Contact avoidance is calculated as the centered 3-week rolling average of raw proportions of respondents avoiding contact while predicted contact is based only on national incidence from May 2021 through the end of the survey in June 2022. Mean Pearson's correlation is -0.40 (sd = 0.23).

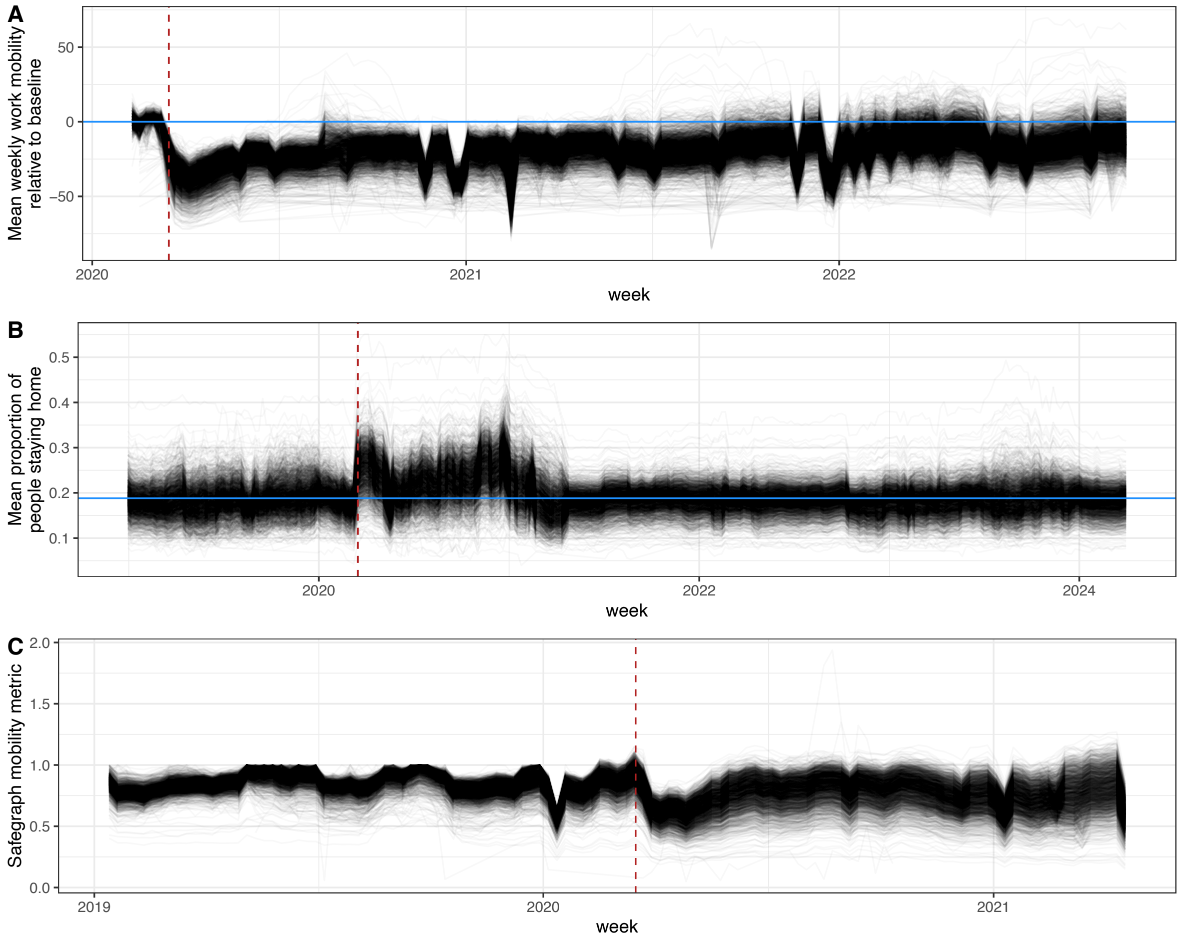


**Figure S54. Mobility time series show precipitous drop in mobility in March 2020 and return to near normal levels towards the end of 2022.** (A) Mean weekly work mobility relative to baseline from Google [76]. (B) Bureau of Transportation Statistics mean proportion of people staying home [77]. (C) Safegraph Social Distancing Dataset mobility metric [48].

**Table S1. Demographic, social, and spatial characteristics of respondents before and after reweighting compared to Census data.**

| Characteristic | Num. responses | Unweighted % | Weighted[1] % | Census[2] % |
|---|---|---|---|---|
| **Total** | 10,680,677 | | | |
| **Age** | | | | |
| 18-24 | 562,590 | 5.3 | 7.1 | 11.87 |
| 25-34 | 1,681,146 | 15.7 | 22 | 17.75 |
| 35-44 | 1,968,526 | 18.4 | 16 | 16.61 |
| 45-54 | 1,941,949 | 18.2 | 16 | 16.29 |
| 55-64 | 2,122,542 | 19.9 | 18 | 16.76 |
| 65-74 | 1,782,728 | 16.7 | 15 | 12.36 |
| $\geq$ 75 | 621,196 | 5.8 | 5.4 | 8.34 |
| **Gender** | | | | |
| Man | 3,517,266 | 32.9 | 48 | 49.01 |
| Woman | 7,163,411 | 67.1 | 52 | 50.99 |
| **Race/ethnicity[3]** | | | | |
| Hispanic | 676,968 | 10.7 | 13 | 16.43 |
| White | 4,885,169 | 77.1 | 61 | 62.40 |
| Asian | 128,916 | 2.0 | 4.8 | 5.88 |
| Black/African American | 361,854 | 5.7 | 11 | 12.20 |
| American Indian/Alaska Native | 57,853 | 0.9 | 1.6 | 0.78 |
| Native Hawaiian/Pacific Islander | 12,269 | 0.2 | 0.4 | 0.18 |
| Multiple/Other Race | 211,513 | 3.3 | 7.5 | 10.84 |
| **Education[4]** | | | | |
| Less than high school | 155,678 | 2.7 | 2.9 | 11.23 |
| High school or equivalent | 936,470 | 16.5 | 17 | 27.25 |
| Some college, no degree | 1,529,925 | 27.0 | 27 | 22.08 |
| Associate's degree | 717,845 | 12.7 | 12 | 8.34 |
| Bachelor's degree | 1,641,593 | 29.0 | 29 | 19.47 |
| Graduate or professional degree | 683,459 | 12.1 | 12 | 11.64 |
| Missing | 4,950,418 | | | |
| **Census region** | | | | |
| Midwest | 2,605,381 | 24.4 | 24 | 20.66 |
| Northeast | 1,856,746 | 17.4 | 17 | 17.23 |
| South | 3,901,439 | 36.5 | 36 | 37.65 |
| West | 2,317,111 | 21.7 | 22 | 23.46 |
| **Household size** | | | | |
| 1 | 1,681,964 | 15.7 | 15 | 11.34 |
| 2 | 3,753,965 | 35.1 | 35 | 27.34 |
| 3 | 1,887,160 | 17.7 | 18 | 18.81 |
| 4 | 1,777,194 | 16.6 | 17 | 20.77 |
| 5 | 818,765 | 7.7 | 7.7 | 12.09 |
| 6 | 413,587 | 3.9 | 3.9 | 5.55 |
| $\geq$7 | 348,042 | 3.3 | 3.4 | 4.10 |

[1] All estimates are from the full survey period (June 2020 – April 2021) and weighted proportions are derived via raking on age and gender only, unless otherwise noted. Percentages may not sum to exactly 100 because responses with more than 72 contacts have been excluded from this table but were included in the raking scheme. Raking is performed at the county-week scale whereas these values are national estimates.
[2] Census percentages are from the 2021 5-year ACS survey, limited to those 18 and older.
[3] All race/ethnicities are Non-Hispanic, unless otherwise noted. Race/ethnicity data was not collected from respondents until September 2020. These race/ethnicity estimates concern only the responses from September 2020 – April 2021 where race/ethnicity was recorded in the survey and the data are raked on race, in addition to age and gender. Therefore, all individuals with missing race responses ($\approx$4% of the valid sample) are not included.
[4] Education is only available starting in September 2020 but these statistics capture the full survey period; thus, many of the missing responses are a result of the question not being on the survey.

Table S2. **Based on mean adjusted-$R^2$, models using objective risk explain equal or more variation in contact patterns than those using subjective risk.** Proportion worried represents the centered 3 week rolling average proportion of CTIS respondents reporting that they are somewhat or very worried about someone in their immediate family becoming seriously ill from COVID-19. Models are run separately for each county for the period from October 1, 2020 through April 24, 2021 and the value provided is the mean across all county models. Models missing reported worry for any week are excluded; this leaves 2,115 counties in the analysis.

| **Predictor scale** | **Incident cases** Mean adj-$R^2$ (sd) | **Proportion worried** Mean adj-$R^2$ (sd) |
|---|---|---|
| County | 0.639 (0.23) | 0.434 (0.24) |
| State | 0.756 (0.16) | 0.742 (0.13) |
| National | 0.861 (0.08) | 0.722 (0.12) |

### Supplemental References


62. Meta. User Guide for the COVID-19 Trends and Impact Survey Weights; 2022. Version 1.

63. Salomon JA, Reinhart A, Bilinski A, Chua EJ, La Motte-Kerr W, Rönn MM, et al. The US COVID-19 Trends and Impact Survey: Continuous Real-Time Measurement of COVID-19 Symptoms, Risks, Protective Behaviors, Testing, and Vaccination. Proceedings of the National Academy of Sciences. 2021 Dec;118(51):e2111454118.

64. Pasek J. Anesrake: ANES Raking Implementation; 2018. https://CRAN.R-project.org/package=anesrake.

65. Wood SN. Generalized Additive Models: An Introduction with R. Second edition ed. Chapman & Hall/CRC Texts in Statistical Science. Boca Raton: CRC Press/Taylor & Francis Group; 2017.

66. Mossong J, Hens N, Jit M, Beutels P, Auranen K, Mikolajczyk R, et al. Social Contacts and Mixing Patterns Relevant to the Spread of Infectious Diseases. PLOS Medicine. 2008 Mar;5(3):e74.

67. DeStefano F, Haber M, Currivan D, Farris T, Burrus B, Stone-Wiggins B, et al. Factors Associated with Social Contacts in Four Communities during the 2007–2008 Influenza Season. Epidemiology & Infection. 2011 Aug;139(8):1181–1190.

68. Feehan DM, Cobb C. Using an Online Sample to Estimate the Size of an Offline Population. Demography. 2019 Dec;56(6):2377–2392.

69. Nelson KN, Siegler AJ, Sullivan PS, Bradley H, Hall E, Luisi N, et al. Nationally Representative Social Contact Patterns among U.S. Adults, August 2020-April 2021. Epidemics. 2022 Sep;40:100605.

70. Dorélien AM, Venkateswaran N, Deng J, Searle K, Enns E, Alarcon Espinoza G, et al. Quantifying Social Contact Patterns in Minnesota during Stay-at-Home Social Distancing Order. BMC Infectious Diseases. 2023 May;23(1):324.

71. Feehan DM, Mahmud AS. Quantifying Population Contact Patterns in the United States during the COVID-19 Pandemic. Nature Communications. 2021 Feb;12(1):893.

72. Kiti MC, Aguolu OG, Zelaya A, Chen HY, Ahmed N, Batross J, et al. Changing Social Contact Patterns among US Workers during the COVID-19 Pandemic: April 2020 to December 2021. Epidemics. 2023 Dec;45:100727.

73. Breen CF, Mahmud AS, Feehan DM. Novel Estimates Reveal Subnational Heterogeneities in Disease-Relevant Contact Patterns in the United States. PLOS Computational Biology. 2022 Dec;18(12):e1010742.

74. Crawford FW, Jones SA, Cartter M, Dean SG, Warren JL, Li ZR, et al. Impact of Close Interpersonal Contact on COVID-19 Incidence: Evidence from 1 Year of Mobile Device Data. Science Advances. 2022 Jan;8(1):eabi5499.

75. Hale T, Angrist N, Goldszmidt R, Kira B, Petherick A, Phillips T, et al. A Global Panel Database of Pandemic Policies (Oxford COVID-19 Government Response Tracker). Nature Human Behaviour. 2021 Apr;5(4):529–538.

76. Google. COVID-19 Community Mobility Report; 2021. https://www.google.com/covid19/mobility?hl=en.

77. Daily Travel | Bureau of Transportation Statistics;. https://www.bts.gov/daily-travel.

78. Safegraph. Social Distancing Metrics;. https://docs.safegraph.com/docs/social-distancing-metrics.
